# Analysis of feature influence on Covid-19 Death Rate Per Country Using a Novel Orthogonalization Technique

**DOI:** 10.1101/2021.07.02.21259929

**Authors:** Gaston Gonnet, John Stewart, Joseph Lafleur, Stephen Keith, Mark McLellan, David Jiang-Gorsline, Tim Snider

## Abstract

We have developed a new technique of Feature Importance, a topic of machine learning, to analyze the possible causes of the Covid-19 pandemic based on country data. This new approach works well even when there are many more features than countries and is not affected by high correlation of features. It is inspired by the Gram-Schmidt orthogonalization procedure from linear algebra. We study the number of deaths, which is more reliable than the number of cases at the onset of the pandemic, during Apr/May 2020. This is while countries started taking measures, so more light will be shed on the root causes of the pandemic rather than on its handling.

The analysis is done against a comprehensive list of roughly 3,200 features. We find that globalization is the main contributing cause, followed by calcium intake, economic factors, environmental factors, preventative measures, and others. This analysis was done for 20 different dates and shows that some factors, like calcium, phase in or out over time. We also compute row explainability, i.e. for every country, how much each feature explains the death rate. Finally we also study a series of conditions, e.g. comorbidities, immunization, etc. which have been proposed to explain the pandemic and place them in their proper context. While there are many caveats to this analysis, we believe it sheds light on the possible causes of the Covid-19 pandemic.

**One-Sentence Summary:** We use a novel feature importance technique to find that globalization, followed by calcium intake, economic factors, environmental factors, and some aspects of societal quality are the main country-level data that explain early Covid-19 death rates.

## Main Text

It is easy to see that European and North American countries were disproportionately affected at the onset of Covid-19. This is counter-intuitive, since all these countries have highly functional health systems, a well educated and nourished population, and are states of law. Solid medical services, sanitation, ample living space, good public transportation, well organized cities, etc. are all arguments for having the disease spread more slowly or not at all, yet the opposite happened.

We propose to analyze this contradiction with the help of machine learning, in particular with the use of feature importance (FI) (e.g. Shapley, SHAP values *(2)*, permutation importance *(3-5)*, partial derivatives, sensitivity analysis *(6-8)*, and several other indices that we have developed).^1^ In the end, due to the peculiarities of the data, we settled on a new method that we have developed for this purpose called orthogonalization. The orthogonalization method is inspired by the Gram-Schmidt *(1)* procedure from linear algebra. It finds the most significant feature for each iteration. By most significant we mean the feature that decreases the *L*^2^norm by the most. (See below).

We aggregated around 3,500 values as features for each country. This analysis should find de novo which are the features that affect positively or negatively the mortality of the disease. These results are conjectures on the causes and need to be verified explicitly with standard medical/biological procedures. Although the relations found could be correlation, causation, proxies, etc., they still contribute to understanding what matters and what does not matter for the mortality of the disease.

We study the number of deaths per population, which is more reliable than the number of cases (see Caveats, 5). We collected five 4-day groups, starting on April 15th, every 10 days until May 28th. We call each date/problem a dataset. These dates are at the onset of the pandemic and before most countries started taking preventative measures, so more light will be shed on the root causes of the pandemic rather than on its handling. Analyzing different dates is crucial for understanding the stability of the results and for discovering trends.

We also compute what is called row explainability, i.e. for every row (country), how much each feature contributes to the target value. These are represented as coloured maps of the world for each feature.

Finally we also study a series of conditions, e.g. comorbidities, immunization, environmental factors, etc. These conditions have been proposed in the literature or in the media as influencing the pandemic. In our study, we compared them against the selected features, and measured the difference of their performances.

## Results: features affecting mortality rates

We ran the orthogonalization algorithm for a maximum of 8 iterations on the 20 datasets. The results of the approximation are quite good - on the average the total *R*^2^is 82.98% (the *L*^2^norm of the errors was reduced by 82.98% from baseline using ML models trained on most prominent features). The results of orthogonalization can be found in the dataset-iteration appendix. From the 20 datasets only 69 features are significant. For each feature we aggregated its importance from all of the 20 datasets. The complete list of FI results with explanations can be found in the feature importance appendix. The features can be thematically grouped as:

| Feature group | importance |
| --- | --- |
| Globalisation | 1246.5292 |
| Calcium intake | 158.7874 |
| Economic quality | 133.4908 |
| Environmental values | 59.0442 |
| Societal quality | 27.5681 |
| Measures | 10.5957 |
| Food | 10.2554 |
| Health factors | 9.8594 |
| Other | 3.4833 |

The following figure shows the top 6 feature groups as they evolve during Apr/May. In this case, we added the results for 5 groups of 4 days, equally spaced starting on April 15th ending on May 28th.

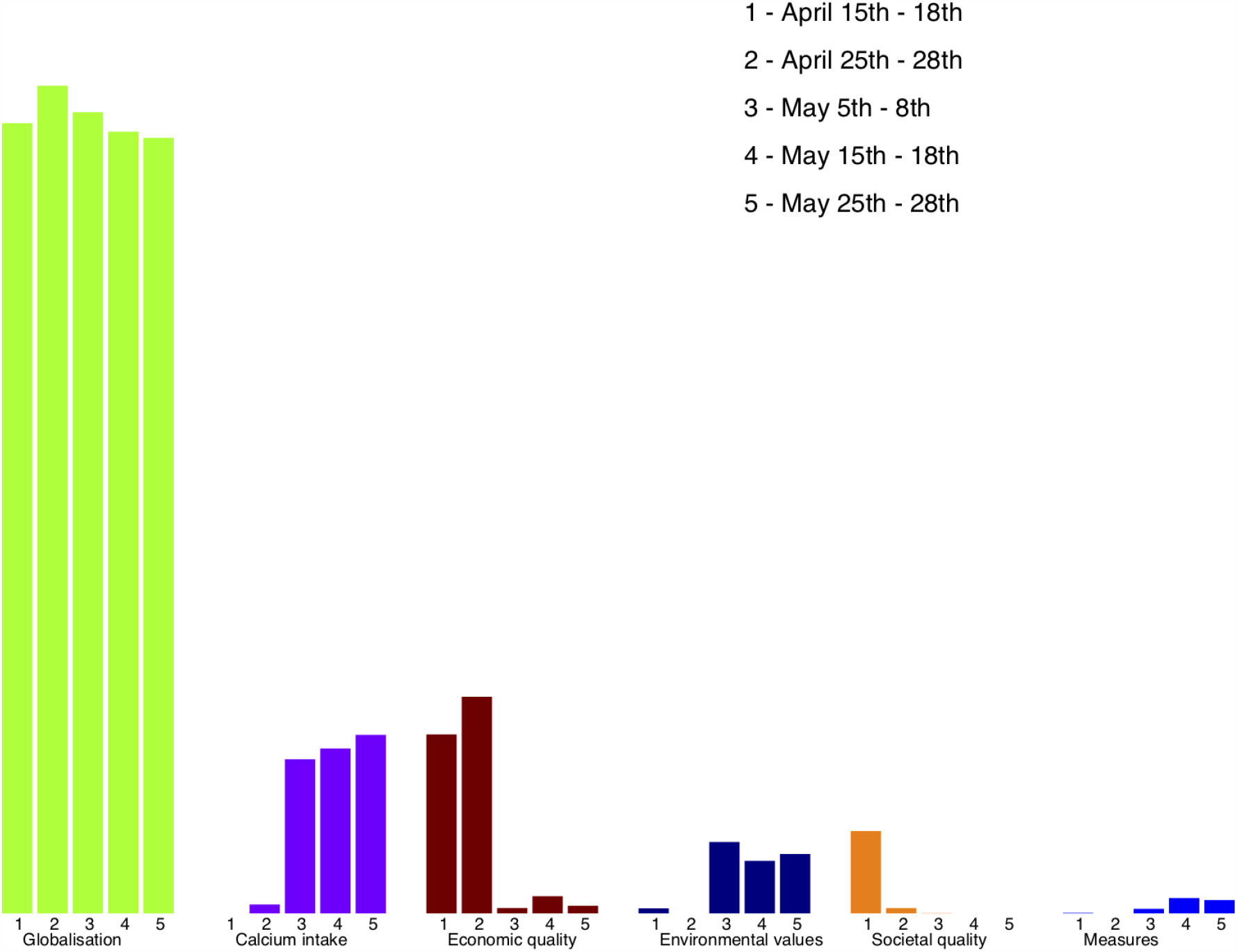

The top group is mainly composed of versions of the index produced by [KOF (ETH Konjunkturforschungsstelle)](https://kof.ethz.ch/prognosen-indikatoren/indikatoren/kof-globalisierungsindex.html#:~:text=Der%20KOF%20Globalisierungsindex%20misst%20die,nach%20Ende%20des%20Kalten%20Krieges) *(37)* which measures various forms of globalisation. It is not surprising at all that globalisation accelerated the dispersion of the virus at earlier times and caused a significant number of casualties. It is quite remarkable how well one single feature contributes to this explanation. Even within the group the top feature collects 98% of the importance. This top feature is defined by KOF as: “Number of international inter-governmental organisations in which a country is a member. International treaties signed between two or more states and ratified by the highest legislative body of each country since 1945. Number of distinct treaty partners of a country with bilateral investment treaties (BITs)”.

The next group is novel - it systematically appears second for May/2020 and hence we believe it is quite solid. Calcium starts appearing at the end of April at the fourth iteration and then phases in as second most significant. It is also very uniform, although we have 72 features related to calcium, the top 2 collect 96% of the importance. The data is taken from the [Global Dietary Database, GDD](https://www.globaldietarydatabase.org/#:~:text=The%20Global%20Dietary%20Database%20(GDD,in%20poor%20and%20vulnerable%20populations)) *(38)*. All the calcium features from GDD are highly correlated with each other. Because of the importance of this result we also analyze it under the conditions section.

Economic quality includes features which cover wealth and good infrastructure. It is not surprising that they appear as significant. This group has 390 features in total, about 20 show up and are difficult to group any further.

Next, there are 271 features related to the environment but one appears clearly at the top. It is Yale’s EPI ([Environmental Performance Index](https://www.researchgate.net/publication/322641392_2018_Environmental_Performance_Index_EPI)) *(39)*. This group becomes significant in May.

Societal quality is a group of features which appears strong in April and then fades out. It has 272 features of which only 7 appear.

Although the health group and the food group have 712 and 570 features respectively, their weight ranks 8th and 9th. It seems from this data that these groups were not an issue at the onset of the pandemic.

Although at this very early stage the data is not strong, we have an indication of some preventative measures that are important. Given the exponential growth of the pandemic, it is crucial to recognize measures which worked earliest. Here is a table of the ones found.

|  |  |  |
| --- | --- | --- |
| Measures |  | 10.5957 |
| OWD Avg. change in visitors to workplaces (all months) | 4.3412 |  |
| OxCGRT H2 Testing policy 3 Open from days 13 to 0 | 1.7421 |  |
| OxCGRT H2 Testing policy 3 Open from days 27 to 14 | 1.4965 |  |
| OWD International travel controls: screening (date) | 0.6006 |  |
| OxCGRT C8 International travel controls 1 screening from days 41 to 28 | 0.5388 |  |
| OxCGRT C2 Workplace closing 2 Require closing/WFH some sectors from days 13 to 0 | 0.4924 |  |
| OxCGRT C7 Restrictions on internal movement 2 Restrictions from days 55 to 42 | 0.4361 |  |
| OxCGRT H2 Testing policy 3 Open from days 41 to 28 | 0.3713 |  |
| OxCGRT C2 Workplace closing 3 Require closing/WFH nonessential from days 13 to 0 | 0.3135 |  |
| OWD Stay at home restrictions: first date any action | 0.1717 |  |
| OxCGRT H2 Testing policy 2 Symptoms from days 13 to 0 | 0.0917 |  |

### Country level analysis

The FI analysis can be done at the row level, which we call *row explainability*. Our rows are the countries, and it is very interesting to find how much each feature explains the mortality rate for each country.

More precisely, for each date and each iteration we are selecting the feature which reduces the *L*^2^ norm the most. The *L*^2^norm is the sum of the squares of the individual targets, one per country.

We define the FI for each country as the reduction of the square of the target (see orthogonalization below).

We have selected two date-iteration pairs to show here. The same selected feature for about the same date gives very similar results. More detailed explanations and further analyses can be found in the row explainability appendix.

For the first analysis, we chose the first iteration for May 15th. The selected feature is *globalization*. Each country is colored in scale with the amount that the square of its target changes. The countries in green are those for which *globalization* explains a significant part of the target, i.e. number of deaths per population. The first feature improves most of the largest deviations, but unfortunately in some cases its predictions worsen the targets, which are shown in red.

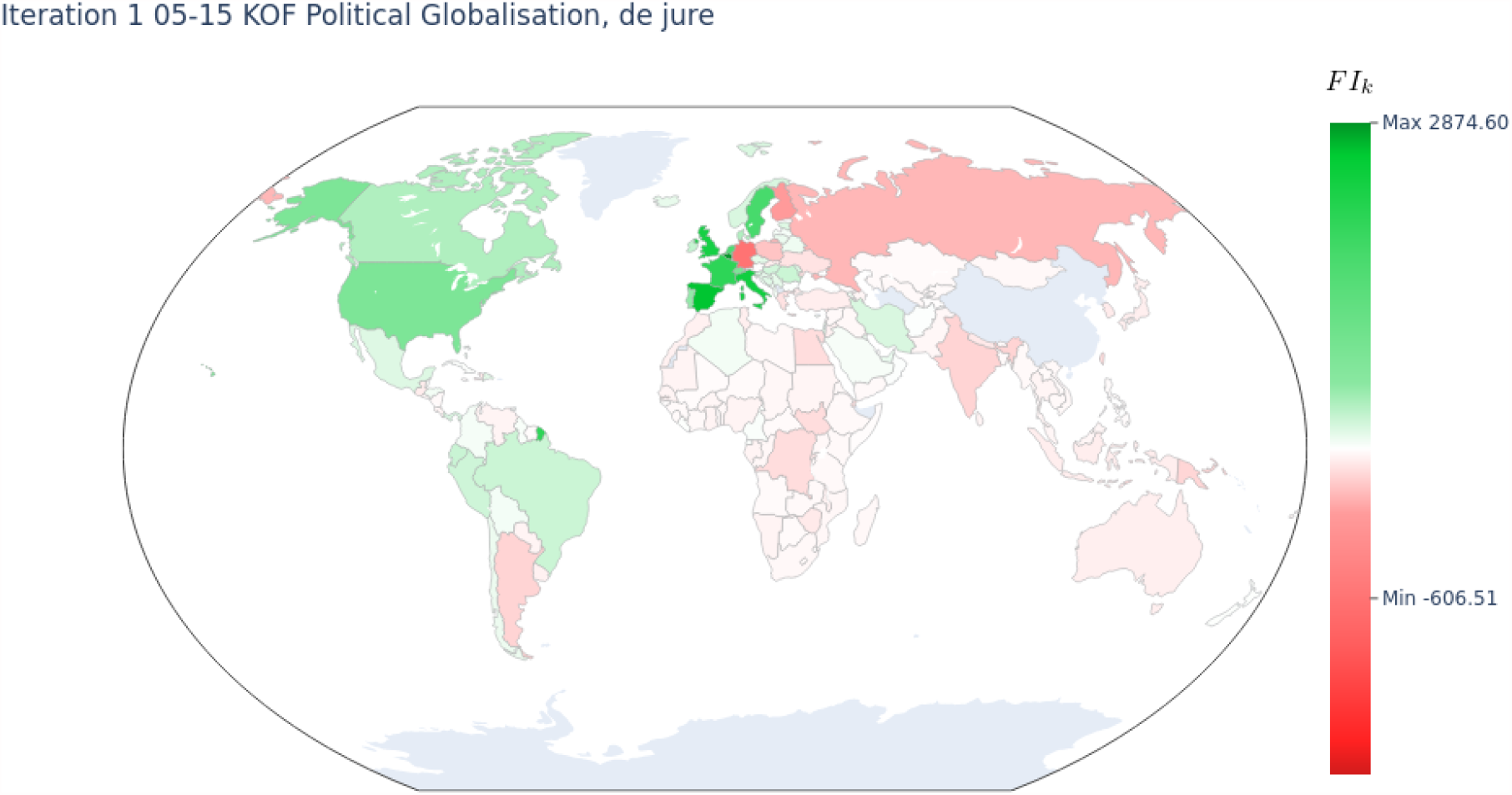

We can see that at this stage the predictions using *globalization* for large parts of Europe and the US improve the targets while for Russia, Germany and other countries, *globalization* worsens them slightly. The latter countries had very low death rates at the onset of the pandemic, while they have a similar level of *globalization* as the former.

The next map shows the second iteration for May 15th, where *calcium* is the selected feature. Here we can see that *calcium* helps explain most of the targets for the northern hemisphere while it worsens them a bit for a few others. It should be remembered, that from all the 3200 features, *calcium* is the one which produces the best results (*L*^2^) for this iteration.

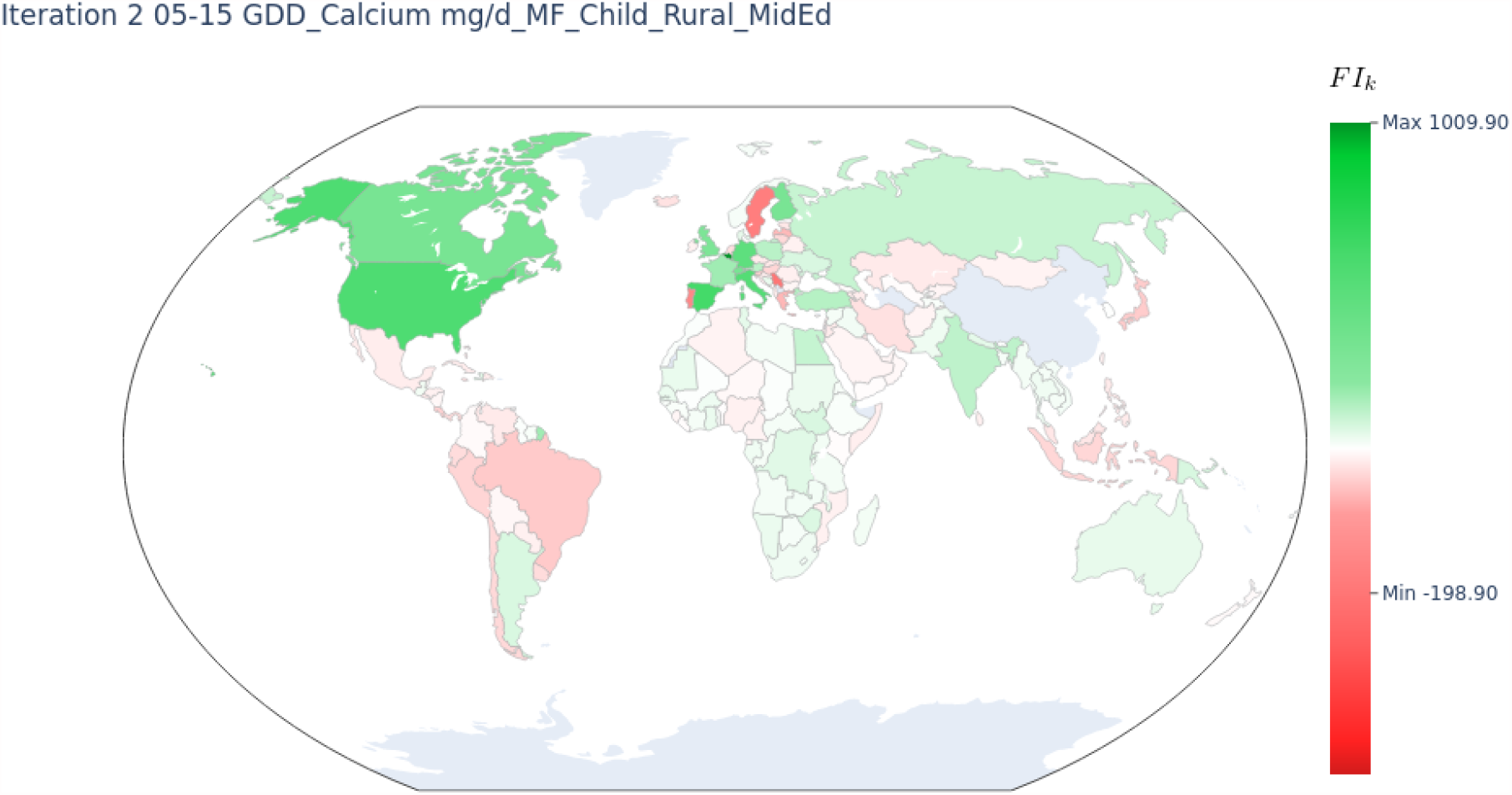

### Results for particular conditions

We call a condition any thematically related set of features which have been the subject of scientific study or have been mentioned extensively by the media, e.g. health risk groups. If some of the components of the condition appear in the orthogonalization process, this is confirmation of their potential contribution. If they do not appear in the process, we want to find where they had the best chances of appearing and how far they were from actually appearing.

This analysis has four possible contributions: (a) which is the most important feature in the condition (b) how it compares to the selected one (c) if the feature suddenly loses importance at some iteration, it is an indication that the selected feature in that iteration is the one which shadows the condition and (d) in some cases, the features in the condition are not significant at all, which provides a strong negative result to the conjecture.

This is done by considering each of the features of the condition at each iteration and comparing its contribution against the selected feature for that iteration. This is already done by the orthogonalization process, as all features are tested at each iteration. In most cases the conditions have some significance but lose against the chosen one. This difference is large (except in one case), which means the conditions analyzed are not significant. This analysis will become clearer with the commented examples below.

The conditions we use are (a) health risk groups, (b) calcium intake, (c) blood type *(23, 43-44)*, (d) immunization (e) inequality (gini index) and (f) preventative measures. The complete information for conditions (c) -(f) are in the corresponding appendices. We show (a) and (b) as examples next.

#### (a) Health risk groups

Several papers have mentioned health risk factors, like hypertension *(9, 29-34)*, diabetes *(9-11)*, obesity *(12-15)*, age *(16)*, etc. *(25-28)* as contributing factors for the fatality rate. In this category we have 276 features which can be found in the feature list for health risks (supplemental materials). The following table shows, for each iteration, which of the risk groups is the most significant and quantifies its contribution in terms of *R*^2^

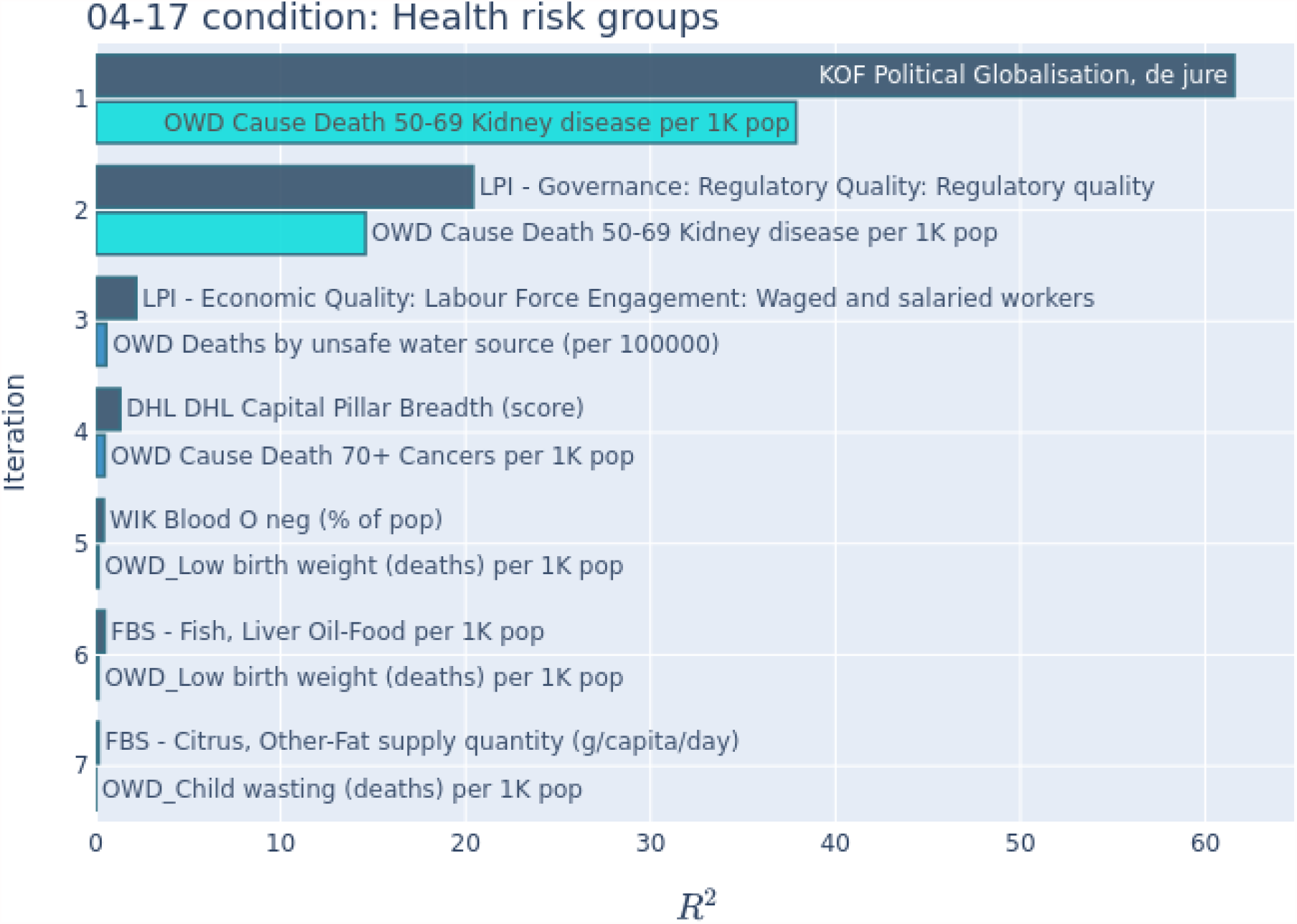

For this dataset we can see that *kidney disease (17, 18)* is the most important feature in the condition group. For the first two iterations it is only 61% and 71% of the *R*^2^of the selected feature, which we consider not significant. The third iteration shows a very large drop in the absolute and relative *R*^2^. This means that whatever contribution *kidney disease* was having, it was shadowed by *LPI Governance*, as once it is considered, there is no more contribution from *kidney disease*. The 4th to 7th iterations have features with very low *R*^2^, which cannot be considered significant. We also notice a lack of consistency of the features chosen, also a sign of lack of significance.

Consequently, this analysis shows that there is no health risk condition that was significantly affecting the number of deaths at that early stage of the pandemic.

#### (b) Calcium intake

*Calcium (19)* appears as a second selected feature in more than half of the dates. The reason for this analysis is to see where it falls when it is not the selected feature. We select 3 dates to show here. To simplify the results, we do not show iterations where the contribution is *R*^2^ < 0. 5%. We selected 84 features with calcium information which can be found in the feature list for calcium (supplemental materials). The results are:

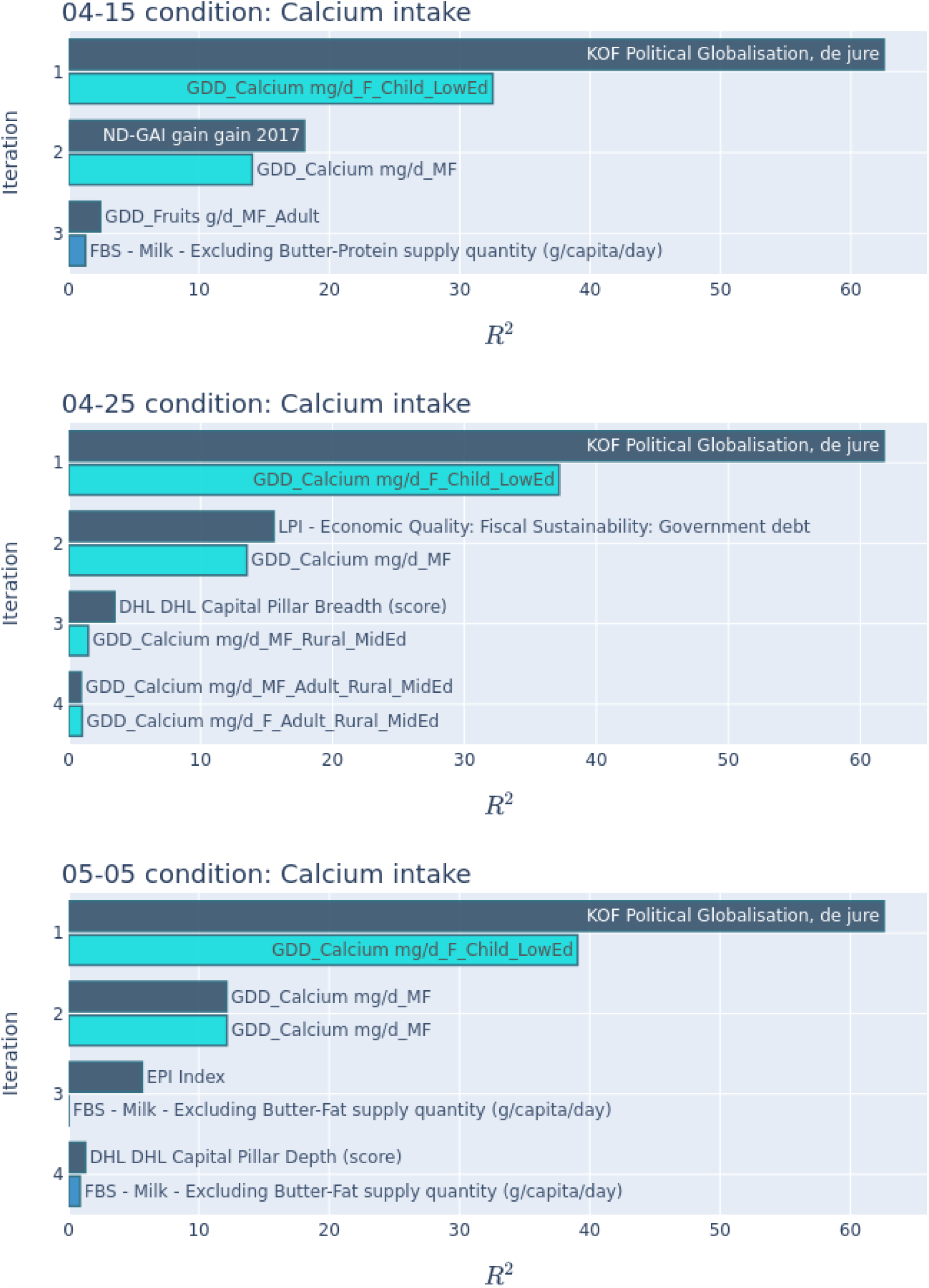

*Calcium* is a strong feature in the second iteration, and as time progresses its relative *R*^2^ goes from 75% to 90% to 100%. This clearly indicates that *calcium* became important gradually. Although the features in the condition include *milk* and *dairy, calcium* is dominant and when *milk* appears is after calcium has been considered. For the first date, it is interesting to see how the *global adaptation index* shadows *calcium*, and for the second date, *economic quality*. This is an indication that these 3 features may be measuring the same phenomenon. We have noticed that all features of *GDD-calcium* are highly correlated, so it is not surprising to see some variation of the best one. The *calcium* data is taken from the [Global Dietary Database](https://www.globaldietarydatabase.org/#:~:text=The%20Global%20Dietary%20Database%20(GDD,in%20poor%20and%20vulnerable%20populations)) *(38)*.

## Methods and materials

### Data gathering

The target variable that we want to study/predict is the number of deaths per population. Being a pandemic for which cases have been reported in more than 200 countries, we will study this target by country. The country granularity is in itself a controversial decision, too coarse for countries like China, Russia or the US, too small for Andorra, San Marino, Malta, etc. We study only countries with population at least 340,000 to eliminate very sparse entries. With these constraints we have information for about 170 countries. We also select multiple dates for the target variable. We take dates during April/May 2020. The reason to take these early dates is to try to identify the causes for the fatality rates. As time went by, treatments became globalised and political decisions diverged, hence it is more difficult to isolate the ground causes.

The target of the study, the number of deaths per population, is readily available from dozens of places, we chose [worldometers](https://www.worldometers.info/coronavirus/#italian-media) *(40)* and [Johns Hopkins](https://coronavirus.jhu.edu/) *(41)*.

For the independent variables, the features, we gathered as much country information as possible. This includes geographic information (size, population, density, climate, coordinates, etc.), demographic information (age distributions, ethnic origins, religions, life expectancy, education, rural vs urban, etc.), medical information (hospitals/doctors per population, prevalence of some diseases, immunization, etc.) economic/development information, education information, and indices (World bank, prosperity, transparency, environmental, inequality, etc.). We have collected more than 3500 features. We include these features when they have data for at least 37.5% of the countries. There are also several other criteria for eliminating features which ends up leaving us with about 3220 features. The sources are very diverse, the UN, WHO, WB, scientific papers, wikipedia and various academic and government agencies are high in the list. The entire list of features with several of their numerical properties can be found in the feature list in the supplemental materials. The values for all features and countries for May 28th can be found in the data in supplemental materials. There are small variations on the number of features and the number of countries for different dates due to missing data.

### Many datasets

We call a dataset all the data needed to make an ML predictor or an FI analysis. This means all the features for a particular set of countries and a target variable. In statistical terms, a dataset is the matrix of independent variables and the target vector. We produce 20 datasets by selecting targets from different dates. We have five groups of 4 dates each, starting on April 15th and every 10 days until May 28th. The datasets have between 3264 and 3295 features. Having many datasets allows us to gain confidence in the results, knowledge about how stable these are and how they evolve in time.

It is known that countries were reporting their data with some random delays. To mimic this we chose the target value (deaths) from a random date, (Uniform(−1,+1) from the date for the dataset) for each country.

As is customary in many branches of science, we use negative controls *(24)*. Details about the negative controls can be found in the dataset iteration appendix.

The diversity forced on targets, plus studying contiguous days, plus the use of negative controls, plus features that appear several times give us confidence in the results. On the other hand, features which appear in the last iterations and in only one dataset are probably less trustworthy.

### Hyperflat problem

With this data, we have a problem with around 170 rows and 3200 columns. The data matrix is extremely flat, and the problem is totally overdetermined. This places quite a bit of stress on the ML algorithms, most of which have not been designed for this shape of problem, rather the reverse. Hyperflat problems also appear in bioinformatics, e.g. gene expression data is of the same shape (tens of thousands of genes, a few tens/hundreds of patients).

Additionally, we have to contend with many features which are highly correlated.

### Machine learning training/testing

The problem, as stated, is a regression problem for which we have 16 methods available to build predictors. The predictors are judged on the accuracy of the prediction, in this case with the *L*^2^ norm of the resulting errors. For each method we try about 10 to 100 combinations of hyperparameters. The predictors are evaluated with cross-validation *(20-22, 35-36)* (randomly selected for each trial) and sufficiently many times to be statistically significant. Since the computation is quite formidable, we use ideas of exploration/exploitation to make this more effective. Roughly speaking this means we place most of the effort in the most promising combinations. From this, rather long, procedure we obtain the best method with its best hyperparameters. We have not kept accurate data on the time spent computing except for the last complete run which used 2.1M core-hours. All in all we estimate we used more than 7M core-hours.

### Feature importance (FI)

Predictors are the main goal of ML. It is important that they are as accurate as possible. Predictors use features as input, but do not tell us how important each feature is for the prediction. What we want in this context is to find which features they use to make the prediction. This is called feature importance, (FI) i.e. which and how much each feature is contributing to determine the answer. Since we have about 3200 features, the ones most important to determine the number of deaths are extremely interesting. This is not a proof by itself, but a strong indication, after which the medical profession can accept or reject the finding.

A feature which shows no importance is likely to be not relevant. While extremely contrived examples could be very important and still show no FI, this is unlikely to happen. In other words, the positives (high FI) are subject to further study. The negatives can be ignored.

FI should not be confused with correlation. FI finds which features allow us to conclude or explain the target. It may be a positive correlation, or it may be a negative correlation or it could be some other, more complex, dependency. It means that some of the ML algorithms are able to extract information from it to predict the target value.

### Orthogonalization

The algorithm that we use for FI is reminiscent of the Gram-Schmidt orthogonalization process adapted to ML. We have developed this algorithm for this particular problem. In simple terms, it works in consecutive iterations as follows. At the start of each iteration we select the feature which, when used with the previously selected features, produces the best predictor. In this case, *best* means reducing the *L*^2^ norm of the error by the most. Once the best feature has been selected, a new target is computed; the previous target minus the contribution explained by the selected feature. And the next iteration can start. This gives us a sequence of features usually in decreasing order of importance and with a numerical value (the relative reduction of the *L*^2^, or *R*^2^) indicating its importance. This is a high level description of the procedure, the subject of this algorithm is reported in a forthcoming computer science paper.

### Instability of orthogonalization

The orthogonalization algorithm, with the Covid-19 data, is not completely stable. If we were to rerun a dataset, typically the first few iterations give the same results, but as the process continues, the inherent randomness of the predictors together with the highly correlated features cause larger and larger variation on the selected features. Often this just means different order of selection, but sometimes different features are selected. This is not inherently wrong. E.g. that calcium intake is selected in one run and milk consumption is selected in another (both highly correlated) may depend on random choices. This instability, choosing targets with minor date variations and running several contiguous dates becomes a tool to detect overfitting. If a feature appears with very low importance and a single time, it is probably not significant. Vice versa a feature that appears with low importance but repeatedly is likely to be significant.

### Value assigned to importance

The algorithms for FI usually assign a value to the importance. This allows us to rank the features according to their importance. While the numerical value depends on the method, within a method the values are usually continuous. Normally the values are ordered so that higher numerical values mean higher importance. For orthogonalization, the natural measure of importance is the reduction of the *L*^2^norm of the target vector. Each iteration finds the feature which reduces the *L*^2^norm by the most, and the relative value of this reduction (usually called the *R*^2^) becomes the numerical value of its importance.

### Caveats

There is a very long list of caveats to this study. Several of them have been pointed out above. The most important ones are:

1. We find dependencies, which are not proofs. The most significant findings should be analyzed individually.
2. Correlation, causation and proxies. A feature that is found to be significant, could be the cause or could be just correlated to the cause. It is useful to define a proxy of the cause. As a real example, at one point in our study, the number of cash dispensing machines appeared to be an important feature. Now these could be (1) a proxy for economic wealth or (2) a proxy for the amount of cash circulating which helps transmit the virus or (3) not a proxy, the machines themselves which are touched by the general public.
3. The features data is from public sources, external to our group. Most of this data comes from good and reliable sources, like UN, FAO, WHO, WB, CIA, Wikipedia. There is no question that occasionally some of these features contain errors, missing data and other problems. We have deleted some of the data that we found too suspicious, but it is not possible for us to guarantee the quality of all the source data. Some results may be affected by errors in the features.
4. This analysis is only able to find dependencies when the features selected show enough variation for different countries. A feature which is similar for many countries will not be able to show a dependency. E.g. Suppose that gender was a very important factor for Covid-19 fatalities, most countries have about the same ratio of genders, and hence we will not be able to discern this dependency from this data.
5. The country level (geographical) granularity of the analysis is quite arbitrary, although it is difficult to find enough information for other granularities.
6. The target variable is also subject to large errors *(42)*. We had done a similar analysis on the number of cases per population and had to abandon it because the data was clearly less reliable. The fact that most countries produce death certificates and the fact that serious cases of Covid-19 demand hospitalization, makes the mortality rate more reliable. The number of cases, besides reporting problems, is seriously biased by the amount of testing and the asymptomatic cases.
7. Significant parts of this analysis use novel algorithms. While we have confirmed the behaviour of our new algorithms with specially defined tests, there could be algorithmic/computational problems.
8. We are making an effort to make all this data/computations reproducible but this has limitations. Computation time and new algorithms are the main issues.

## Future work

Every person who has seen this analysis has a suggestion on (a) more/new features, (b) different targets, (c) different methods, (d) different dates, etc. These computations are already extremely time consuming. We have to limit this research. Most notably, researchers want to feed the data of the second wave of Covid-19 from the late fall 2020. The analysis of the second wave will be the subject of a forthcoming paper.

## Supporting information

Feature list for blood groups

Feature list for calcium

Feature list for health risks

Feature list for immunization

Feature list for inequality

Feature list for preventative measures

Feature names and properties

Selected features by iteration

## Data Availability

The data is included in the supplementary materials.

https://docs.google.com/spreadsheets/d/e/2PACX-1vTIYyxviw2kTFomkqrIiUDMG-8NP0juPrk-ZPGhCZPpfWO-Gx5BiyS08pk5gto3BSWRjvtRVesxGUzI/pub?output=tsv

## Acknowledgments

The feature data for countries was collected by Mark, John, Stephen and Gaston. Joe suggested the row explainability by country. Gaston and Tim wrote the paper, and together with John and Stephen they edited it. David made several observations to the manuscript. Gaston wrote the programs for the analysis and orthogonalization. John produced most of the graphics. The vast computations were made in our own dedicated computers at ETH (gonnet group) and PolyML and by the Euler supercomputer (ETH Zurich). The authors also wish to acknowledge the contributions of Fred Speckeen and Stephen Benner suggesting the condition groups.

## Appendices

### Appendix I: Dataset Iteration

The results of the orthogonalization procedure on the 20 datasets is shown in a table. Each row describes a dataset with the date, number of countries, number of features and 8 pairs of *R*^2^value and feature name. There are small variations on the number of countries and on the number of features due to missing data. We only include a feature when at least 37.5% of the values are known. The feature names have been abbreviated.

Notice that the first iteration is completely homogeneous (the first feature is always the same). Then there is some variation in the second, third and fourth, but still a noticeable pattern of features phasing in and out. Later iterations are less uniform. This is expected, as the *R*^2^values decrease rapidly.

The table is in the selected Features by Iteration in the supplemental materials.

The entries for *R*^2^which have a yellow/red background are deemed not significant. This is done with our technique of negative controls and requires a more detailed explanation. The level of statistical significance for white is 95%, for yellow 90% and for red below 90%.

#### Significance analysis

We define the null hypothesis for each iteration of orthogonalization as follows: a feature which has been randomly permuted, cannot contribute in any way to explain the target. A feature that is totally independent of the target is part of our null hypothesis.

For the negative controls we use mostly randomly permuted features in addition to a few composed of purely random values (random U(0,1), N(0,1), and randomly shuffled consecutive integers). In a situation where the features may have repeated values and/or missing information, a purely random variable may behave differently. This is unwanted, so the majority of our negative controls are randomly selected features, randomly permuted. We use exactly the same software/procedure to compute the scores of the features and of the negative controls.

At the end of each iteration we want to estimate the probability of the selected feature belonging to the null hypothesis. We can generate an arbitrarily large number of negative controls and see whether the *R*^2^of the best selected feature falls within those of the negative controls. If it falls within the negative controls we have an unbiased estimate of the probability. If it falls outside the range, we estimate its probability with suitable approximations of the distribution. We compute the *R*^2^of at least 500 negative controls. We are interested in 3 cases,

1. White background: Pr{selected best in *H*_0_} < 0.05
2. Yellow background: 0.05 ≤ Pr{selected best in *H*_0_} < 0.1
3. Red background: 0.1 ≤ Pr{selected best in *H*_0_}

We use bootstrapping without replacement (45) with ⅔ of the sample to estimate the consistency of the above decision. If the conclusions are less than 90% consistent we enlarge the number of negative controls. When we reach twice the number of features, about 6500 in our case, we stop enlarging the negative controls. We have inspected all the results and are happy with the approximations.

Finally, we are not testing a single feature, we are testing close to *N* = 3^2^00 of them. So the probability of the best of *N* to be in the null hypothesis has to be adjusted accordingly. Since many features in this problem are highly correlated, we need to correct *N*. This corrected *N*_*corr*_ is an approximation of the dimension of the space spanned by the *N*features. If *p* is the probability of being in the null hypothesis, then:

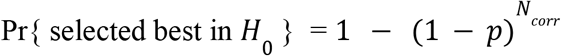

The following graph shows one typical example where we computed 550 negative controls, i.e. a distribution of *H*_0_. The horizontal axis is the *L*^2^ norm (less is better). The vertical axis has no meaning, it is used to spread the points to better visualize them. The vertical bar on the left marks the score of the selected feature. The vertical line on the right marks the initial *L*^2^ (the points to the right mean a worsening of the norm). The continuous jagged line is the cumulative distribution of *H*_0_. In this case the probability of the selected feature not being better than a random permutation is *p* = 0. 0003758. Considering *N*_corr_, the probability of the best score being in *H* is Pr{sel best in *H*_0_} = 0.05978. This means that we cannot accept this best feature as significant, it is borderline.

To estimate the probability of the best selected feature, *p*, we differentiate between two cases: (1) when the selected feature falls within the negative controls; (2) when the selected feature is better than all negative controls. The first case is a safer estimation; it is an interpolation and the prediction is unbiased regardless of the distribution of *H*_0_. The second case is an extrapolation, and is subject to an assumption about the distribution of *H*_0_.

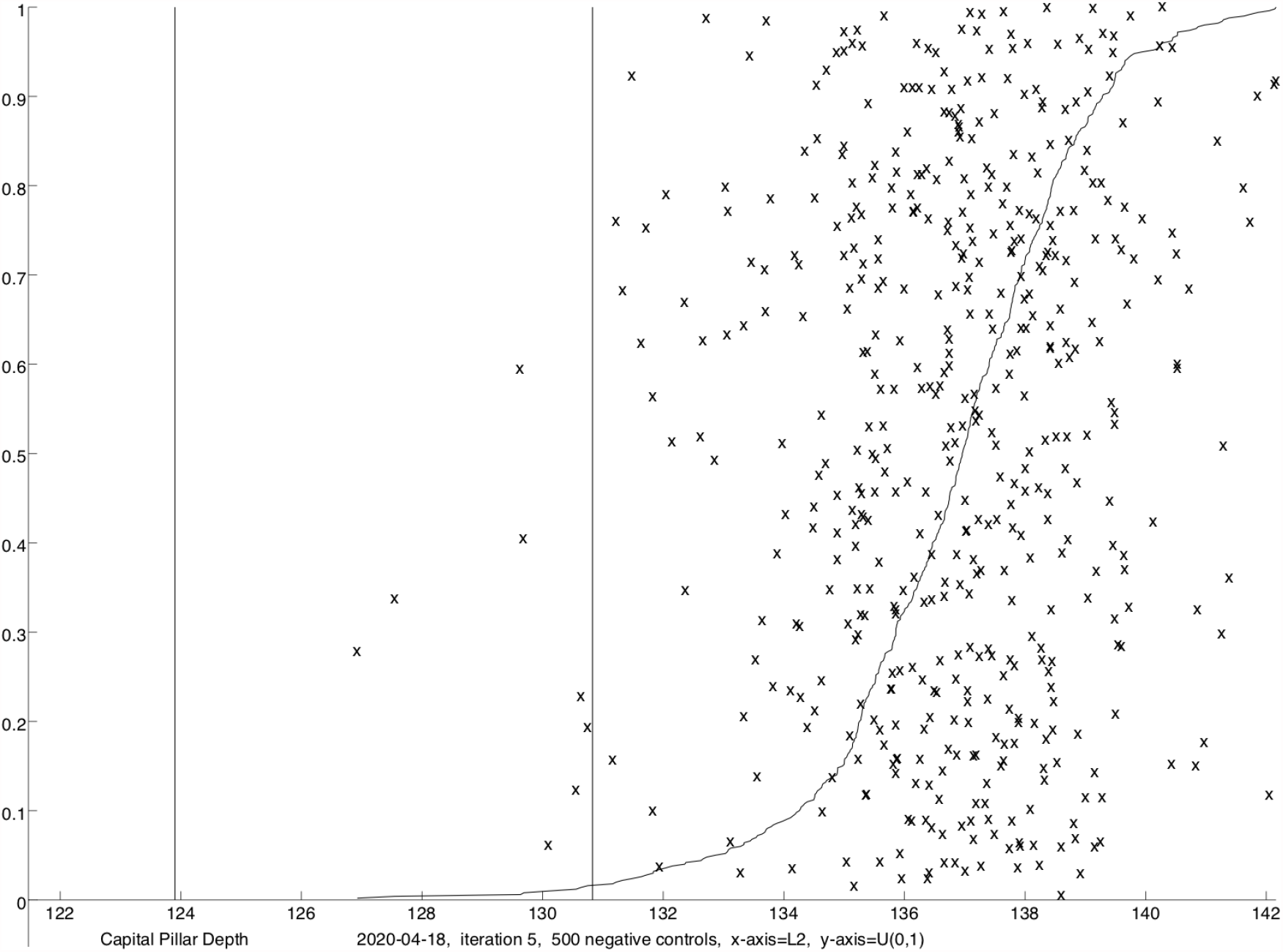

The next graph shows the exponential behaviour of the tail of the *H*_0_ distribution for the same data. This is a graph of the logarithm of the cumulative distribution. For a purely exponential distribution, the cumulative would be a straight line. For a normal distribution, the left part of the graph would approximate a downwards parabola. Hence we try two approximations of the leftmost part of the cumulative: (a) a straight (red) line, choosing the best fit from 6 to 20 points and (b) a parabola, considering from 10 to 40 points and only if the quadratic coefficient is negative. The approximation giving the smallest error is chosen. In this case the linear approximation is chosen because the parabolic approximation has the wrong curvature. The red B indicates the *L*^2^of the selected feature, in the intersection between the extrapolated line and the value of *L*^2^.

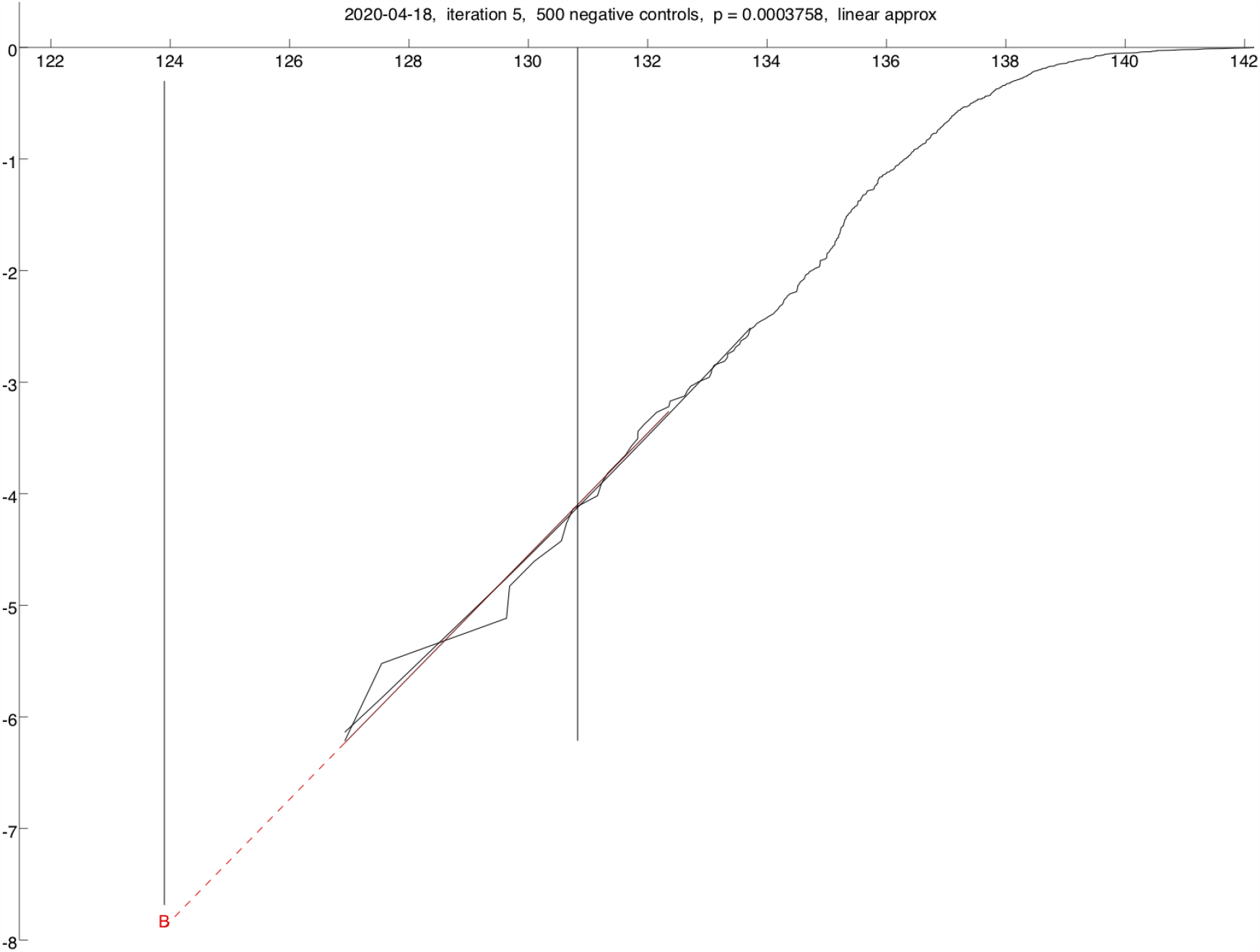

The most important points to fit are the leftmost tail of *H*_0_, as we are extrapolating in that direction. This was the first sample for this case, as the value was so close to the 0.05 boundary, more negative controls were computed.

### Appendix II: Feature Importance

We collect together all the iterations of the 20 datasets to give a global picture of the feature importance. Each individual value is an *R*^2^value, hence a percentage, so they are all of the same magnitude.

For each iteration, the collection of features/values is as follows: (1) one feature is at the top. (2) the same feature may appear several times, usually with different methods/hyperparameters. This is totally expected, e.g. if the top feature is obtained with a random forest, maybe nearest neighbors also gives good results. Only the top feature is considered when it appears many times. (3) any other feature which appears near the top within a reasonable chance of being at the top, is also considered. This is a statistical test: could the expected value of the second feature be greater than the top one with probability > 1/20? (4) when *k* features are possibly close to the top, each one gets 1/*k* of the importance of the top feature (the *R*^2^ value).

This procedure ensures that if two features have about the same importance, both will be counted, and this will not produce a bias. Below there is an example of the output of iteration 4 for April 15th.

We have grouped the features thematically, as in some cases it is clear that a group of features are highly related and correlated. The picture is clearer with the groups. As you can see, in most cases, the top or a couple of the top represent the lion share of the contribution.

Please notice that values have to be compared to 2000 (20 datasets x 100%), so for example an importance of 0.05 is very insignificant. For this reason, features which contribute less than 0.5 are grouped in an “other” category. Also notice that the groups of features have many more entries than shown here, e.g. Environment has over 270 features.

| Feature/group | importance | group total |
| --- | --- | --- |
| **Globalisation** |  | 1246.5292 |
| KOF Political Globalisation de jure | 1221.9791 |  |
| DHL DHL Capital Pillar Depth (score) | 11.74 |  |
| DHL DHL Capital Pillar Breadth (score) | 8.1499 |  |
| KOF Informational Globalisation de jure | 4.1941 |  |
| 3 other features | 0.4661 |  |
| **Calcium intake** |  | 158.7874 |
| GDD Calcium mg/d MF Child Rural MidEd | 111.7066 |  |
| GDD Calcium mg/d MF | 40.2487 |  |
| GDD Calcium mg/d MF Child Rural | 4.0383 |  |
| GDD Calcium mg/d MF Adult Rural MidEd | 1.5592 |  |
| 5 other features | 1.2346 |  |
| **Economic quality** |  | 133.4908 |
| LPI Economic Quality: Fiscal Sustainability: Government debt | 62.7287 |  |
| LPI Governance: Regulatory Quality: Regulatory quality | 35.8991 |  |
| ND GAI gain gain 2017 | 15.877 |  |
| LPI Economic Quality: Fiscal Sustainability | 4.0631 |  |
| WFB Debt external (\$) per 1K pop | 2.287 | |
| LPI Economic Quality: Labour Force Engagement: Waged and salaried workers | 2.1783 |  |
| WFB Telephones fixed lines (lines) per 1K pop | 1.9053 |  |
| WB Foreign direct investment net inflows (BoP current US\$) per 1K pop | 1.8881 | |
| WFB Refined petro product consumption (bbl/day) per 1K pop | 1.4224 |  |
| GII Research and development (R&D) score | 1.2925 |  |
| OWD Debt and contract relief: narrow relief (date) | 1.2244 |  |
| WB Getting Electricity Time (days) | 1.218 |  |
| 9 other features | 1.5068 |  |
| Environmental values |  | 59.0442 |
| EPI Index | 53.6481 |  |
| LPI Natural Environment: Oceans: Overexploitation of fish stocks | 5.0825 |  |
| LPI Natural Environment: Forest Land and Soil | 0.3135 |  |
| Societal quality |  | 27.5681 |
| LPI Living Conditions: Material Resources | 25.3509 |  |
| Human Freedom index ef regulation labor dismissal | 1.1476 |  |
| 5 other features | 1.0695 |  |
| Measures |  | 10.5957 |
| OWD Avg. change in visitors to workplaces (all months) | 4.3412 |  |
| OxCGRT H2 Testing policy 3 Open from days 13 to 0 | 1.7421 |  |
| OxCGRT H2 Testing policy 3 Open from days 27 to 14 | 1.4965 |  |
| OWD International travel controls: screening (date) | 0.6006 |  |
| OxCGRT C8 International travel controls 1 screening from days 41 to 28 | 0.5388 |  |
| 6 other features | 1.8766 |  |
| Food |  | 10.2554 |
| GDD Fruits g/d MF Adult | 2.4451 |  |
| FBS Cocoa Beans and products Fat supply quantity (g/capita/day) | 2.1548 |  |
| **FBS Stimulants Fat supply quantity (g/capita/day)** | 2.1548 |  |
| **FBS Milk Excluding Butter Food supply (kcal/capita/day)** | 1.5619 |  |
| **FBS Coconut Oil Protein supply quantity (g/capita/day)** | 0.7989 |  |
| **FBS Fish Liver Oil Food per 1K pop** | 0.7066 |  |
| **3 other features** | 0.4334 |  |
| **Health factors** |  | 9.8594 |
| **WIK Blood O neg (% of pop)** | 2.7626 |  |
| **LPI Health** | 1.1875 |  |
| **WHO Provincial hospitals (per 100000 pop)** | 1.0454 |  |
| **WIK Blood A pos (% of pop)** | 0.8125 |  |
| **LPI Health: Preventative Interventions: Measles immunisation** | 0.7771 |  |
| **WBD Population 55 to 64 female per 1K pop** | 0.7311 |  |
| **OWD Cause Death All Age Parkinson disease per 1K pop** | 0.6911 |  |
| **WFB Health expenditure (%)** | 0.6606 |  |
| **6 other features** | 1.1918 |  |
| **Other** |  | 3.4833 |
| **UN Geoscheme w49 region 155** | 1.1028 |  |
| **temperate** | 0.7538 |  |
| **7 other features** | 1.6267 |  |

Example of the output of iteration 4 for April 15th of orthogonalization (real output):

~~~
# stored on Sun May 23 11:32:17 2021, Iteration 4
# selected best: Covid-19-Death415, MF: DiscrTest=Kolmogorov, NumberTrees=81, Fraction=0.8, DiffExp=0, Agg=ARand, RdR=1e-05, [759,2814,1095,605]: 90293 +- 521 (4153)
EPI Index, Covid-19-Death415, MF: DiscrTest=Kolmogorov, NumberTrees=81, Fraction=0.8, DiffExp=0, Agg=ARand, RdR=1e-05,
[759,2814,1095,605]: 90293 +- 521 (4153)
EPI Index, Covid-19-Death415, MF: DiscrTest=Kolmogorov, NumberTrees=81, Fraction=0.8, DiffExp=0, Agg=ARand, PCAZ=n ->
round(sqrt(n)), RdR=1e-05, [759,2814,1095,605]: 90332 +- 569 (3601)
WFB Telephones fixed lines (lines) per 1K pop, Covid-19-Death415, MF: DiscrTest=Kolmogorov, NumberTrees=81, Fraction=0.4,
DiffExp=0, Agg=ARand, RdR=1e-05, [759,2814,1095,51]: 92960 +- 245 (3552)
EPI Index, Covid-19-Death415, MF: DiscrTest=Kolmogorov, NumberTrees=81, Fraction=0.4, DiffExp=0, Agg=ARand, PCAZ=n ->
round(sqrt(n)), [759,2814,1095,605]: 93033 +- 556 (3694)
WFB Telephones fixed lines (lines) per 1K pop, Covid-19-Death415, MF: DiscrTest=Kolmogorov, NumberTrees=81, Fraction=0.4,
DiffExp=0, Agg=ARand, PCAZ=n -> round(sqrt(n)), RdR=1e-05, [759,2814,1095,51]: 93160 +- 263 (2951)
EPI Index, Covid-19-Death415, MF: DiscrTest=Kolmogorov, NumberTrees=81, Fraction=0.4, DiffExp=0, Agg=ARand, PCAZ=n ->
round(sqrt(n)), RdR=1e-05, [759,2814,1095,605]: 93929 +- 243 (2678)
… …
~~~

In this case we see that the top feature is the EPI Index which appears twice at the top. In third position we find WFB Telephones fixed lines which is well outside the confidence margin. The rest are different hyperparameters for both features. In this case only the top feature gets the *R*^2^ assigned.

### Appendix III: Row Explainability

#### Formal definition

More precisely, for each date and each iteration we are selecting the feature which reduces the *L*^2^ norm the most. The *L*^2^norm is the sum of the squares of the individual targets, one per country. At the end of iteration *i*

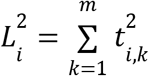

where *t*_*i,k*_ is the target for country *k* at the end of iteration *i* and *t*_0,*k*_ is the initial target for country *k*, the death rate per population. At a given iteration, the targets are predicted by a new ML model. We define a new target as the difference between the initial target for the iteration and the predicted value. I.e. the new target is the unexplained portion of the previous target.

Iteration *i* changes the target *t*_*i*−1,*k*_ → *t*_*i,k*_ with a particular ML model which uses the previous features and the selected feature for iteration *i. model* (*row k*) = *t*_*i*−1,*k*_ − *t*_*i,k*_.

The sequence 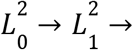 is a decreasing sequence of the errors of the target. The *R*^2^values are 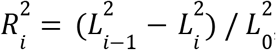, the relative (to initial) decreases in *L*^2^.

We define the FI for each country (*FI*_*k*_) as the normalized reduction of the square of the target 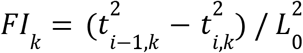. In this way, the sum of the *FI*_k_ for all countries is the 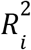.

To fully understand the process, we show one example with full information, that is four graphs, (1) the values of 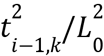, (2) the improvement, i.e. *FI*_*k*_, (3) the resulting values, i.e. 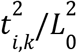 and (4) the values of the selected feature which caused this change.

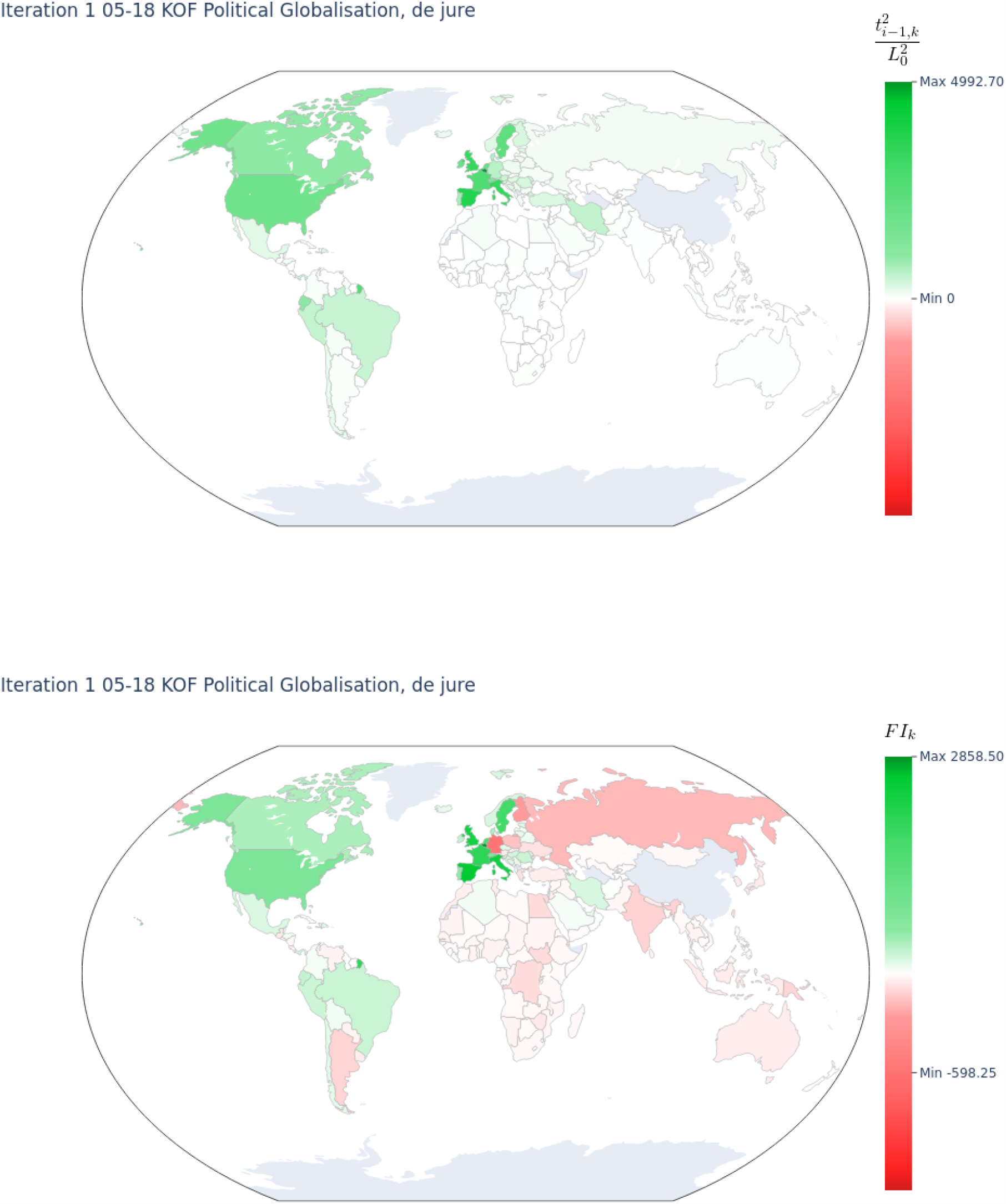

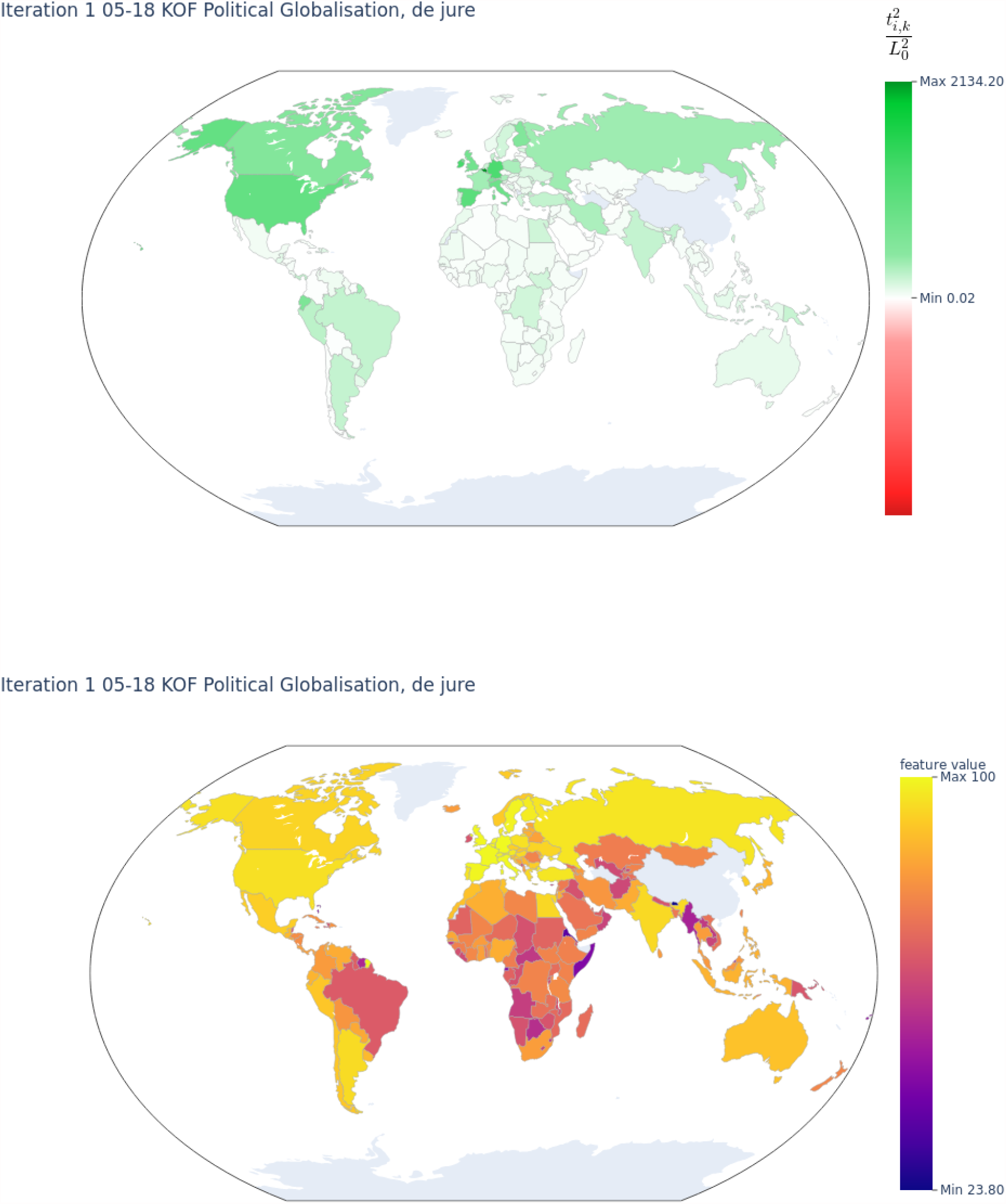

The above are for the first iteration for May 18th. The selected feature is *globalization*. This is too much data, so in general we will show only the second graph, that is *FI*_*k*_ or how much each target was explained.

The following graphs show the first 8 iterations of May 18th. Please notice that as the iterations proceed, the values of *FI*_*k*_ become less significant.

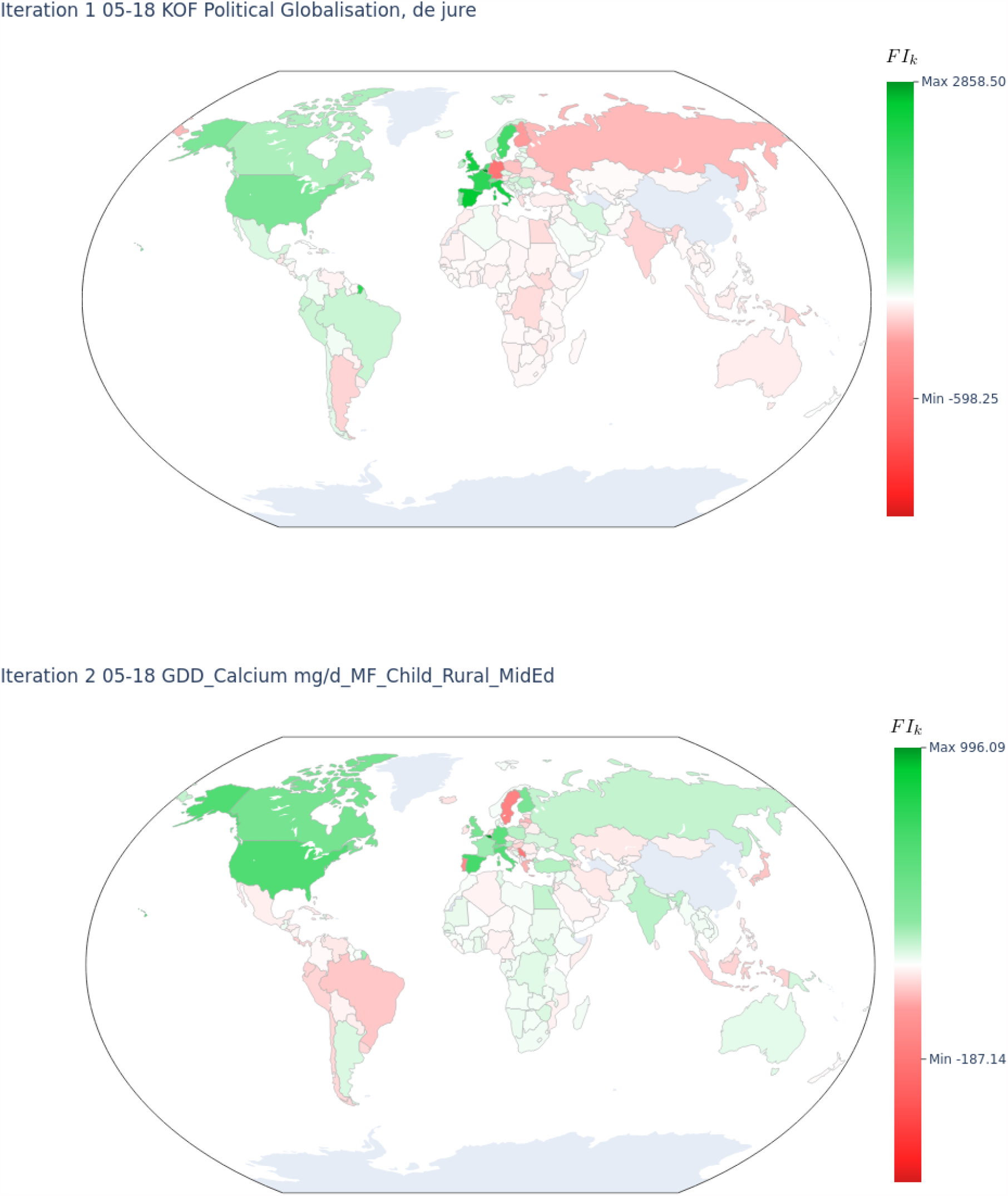

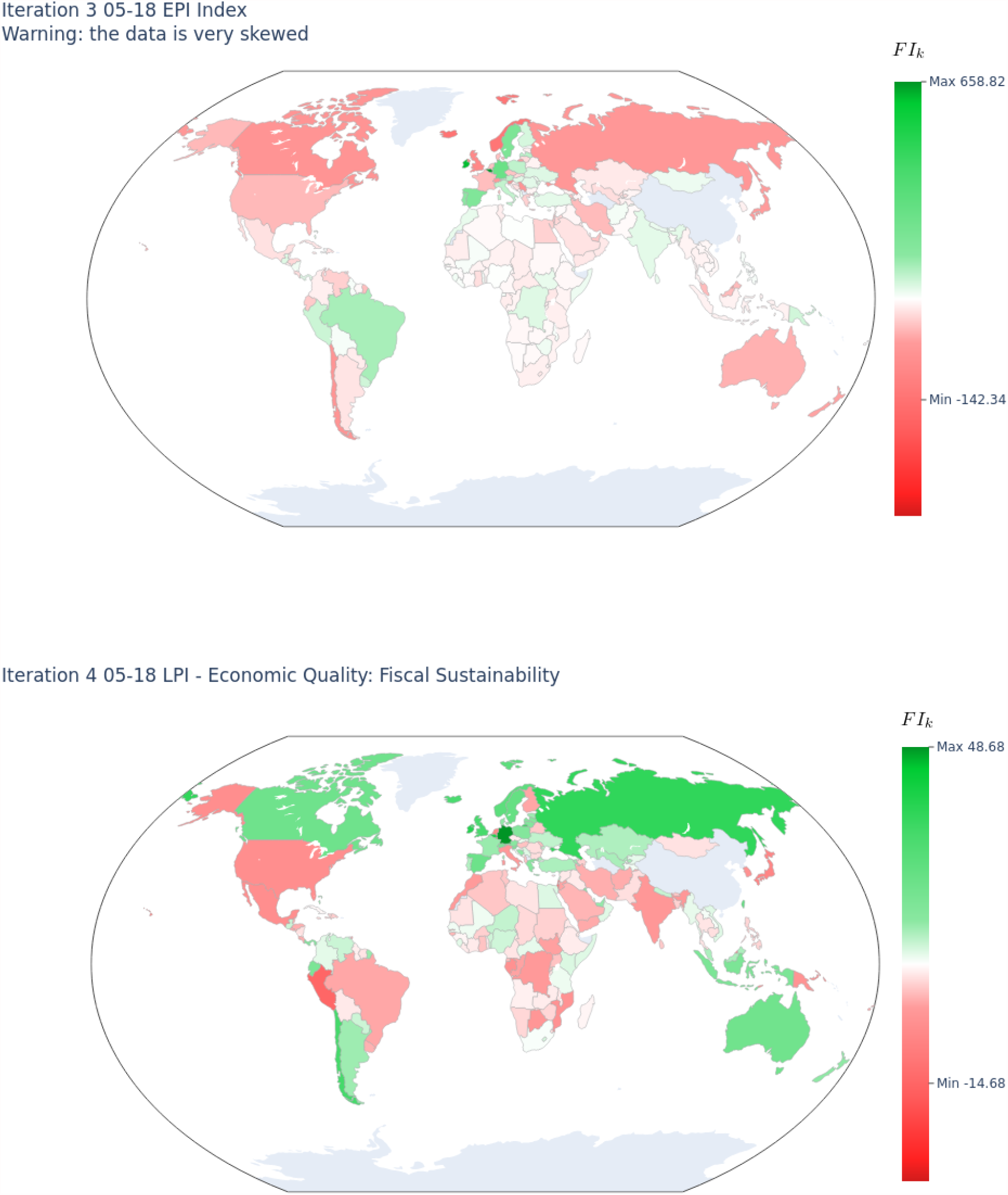

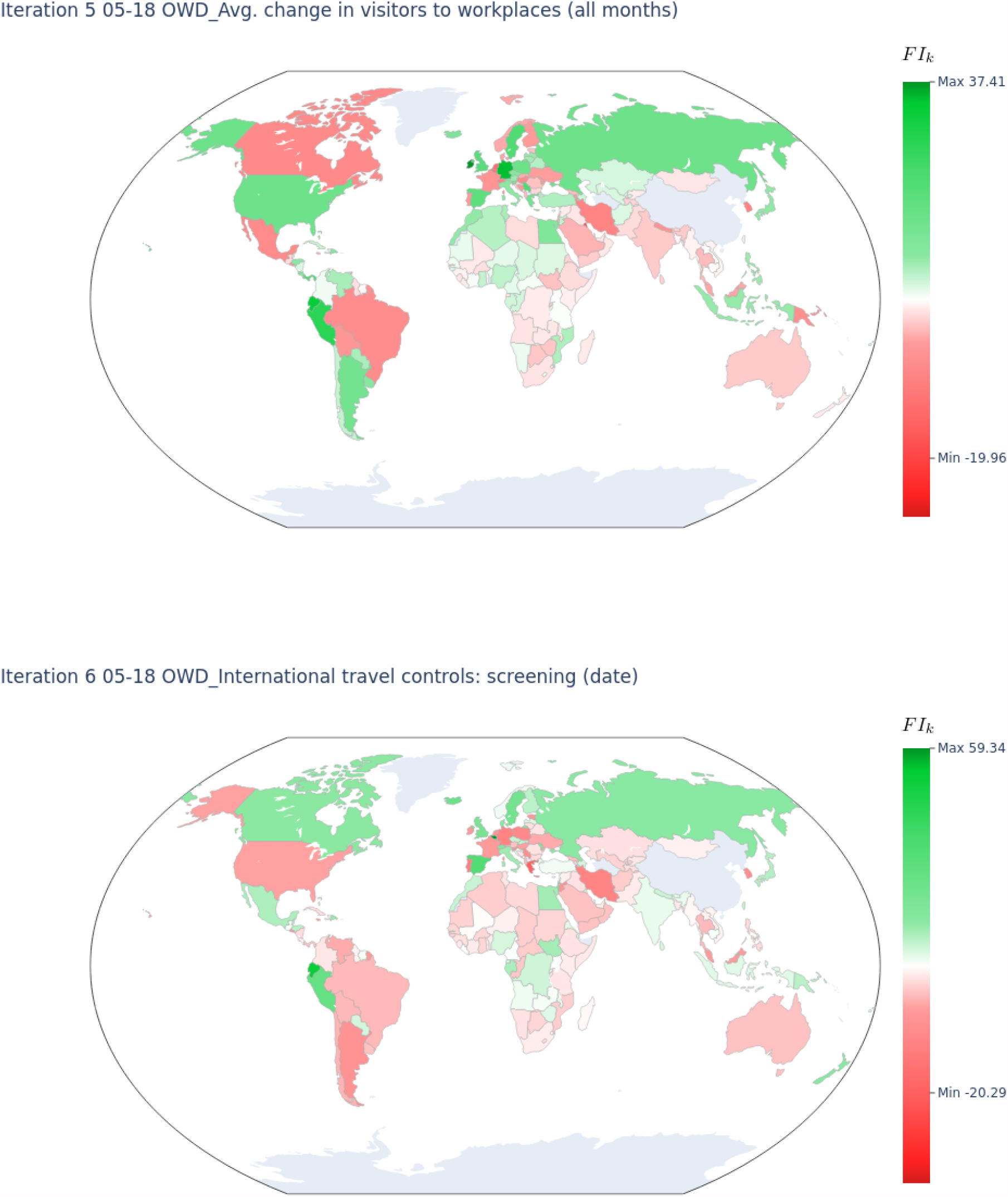

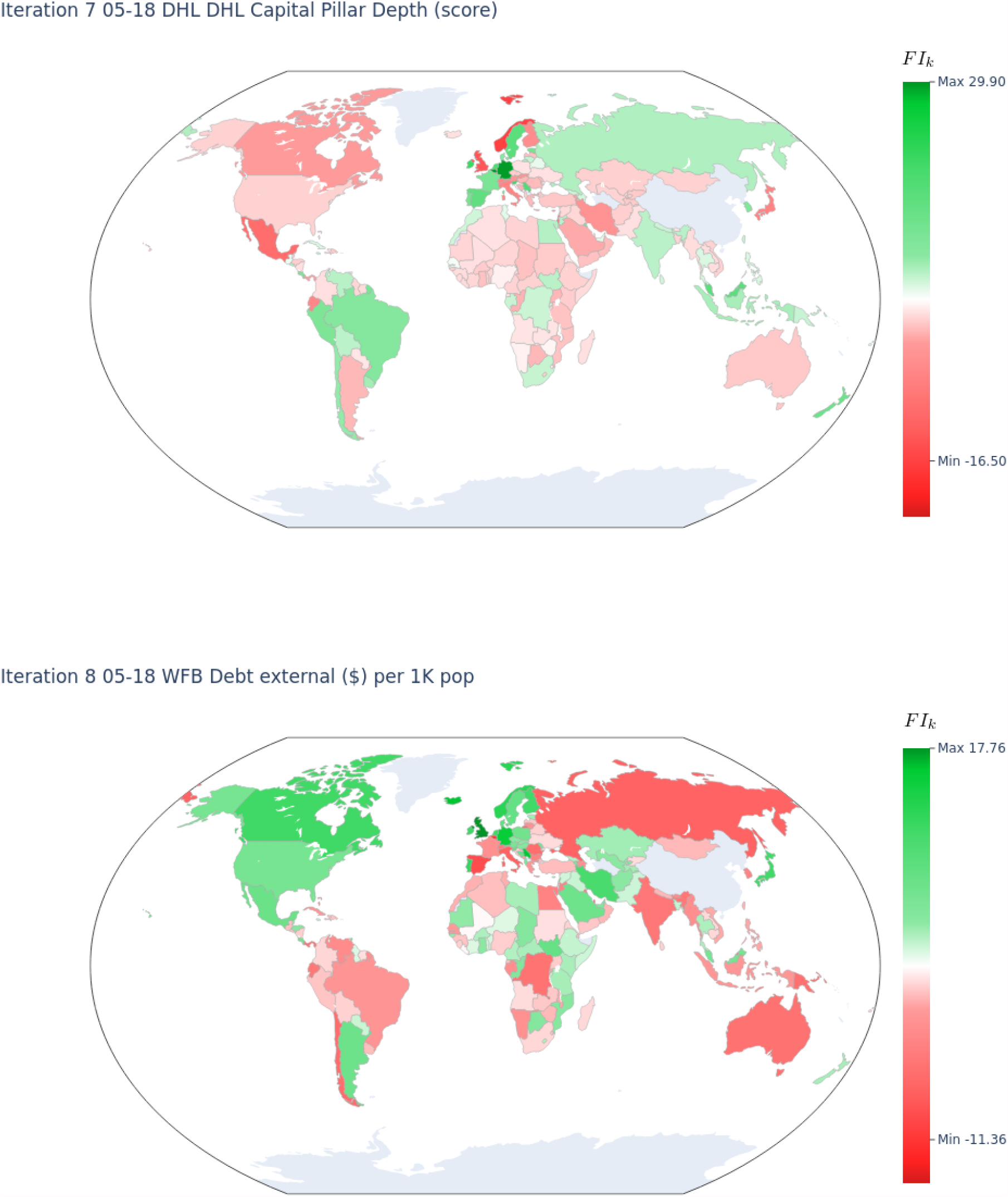

### Appendix IV: Blood Groups

The influence of blood types for Covid-19 infection and mortality rates has been extensively discussed in the literature (23, 43-44). It is natural then to consider them as a condition.

We selected 8 features with blood group information, which can be found in the feature list for blood groups (supplemental materials). The results are:

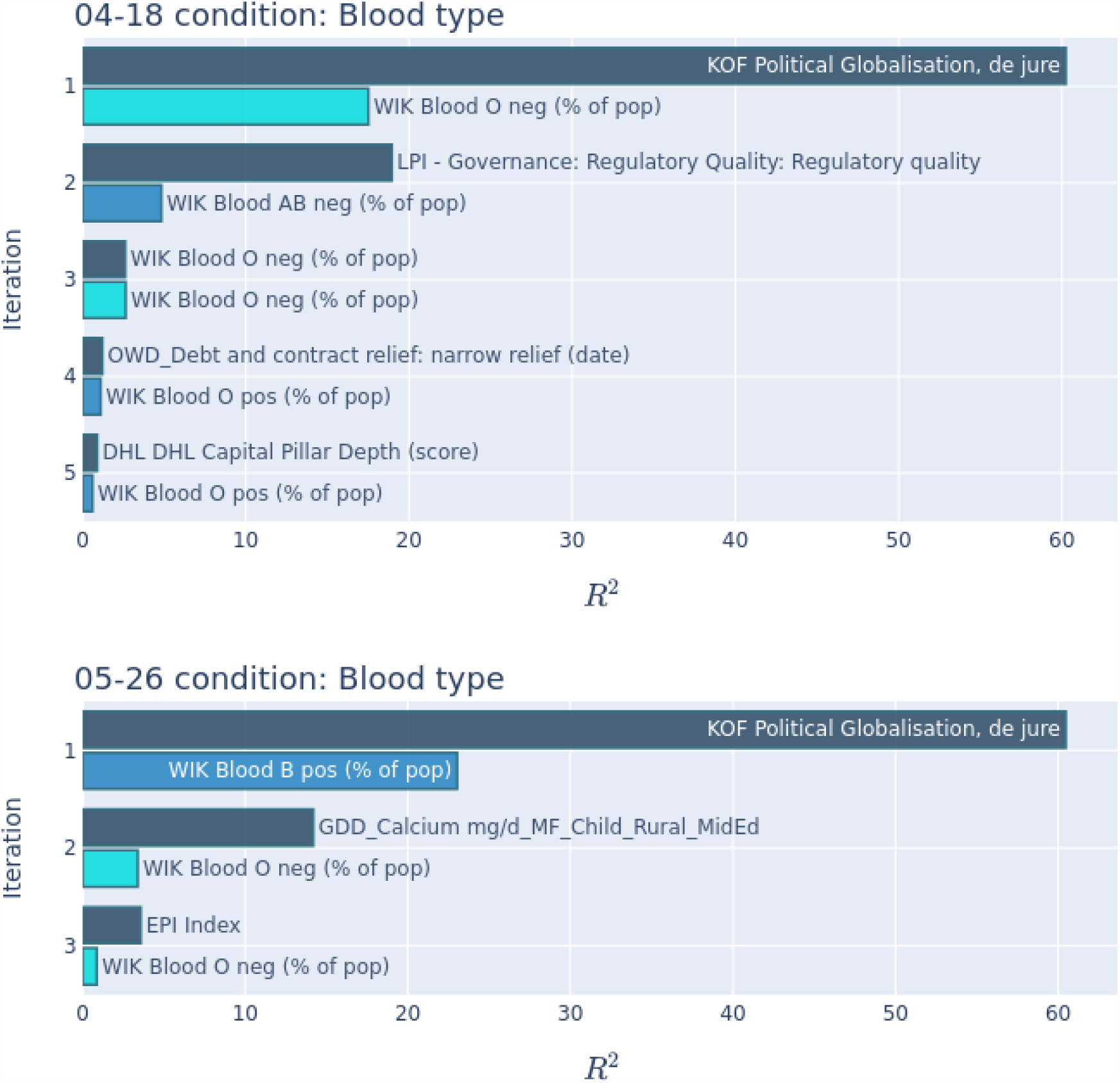

The blood O neg is clearly the dominant feature in this condition. It appears frequently and in one case (04-18) is the selected feature on iteration 3. The analysis shows that the importance of features in this group is minor. Political globalisation is almost 3 times more important (first iteration), and governance and calcium both 4 times more important in their respective iterations. Its importance continues to decrease in the following iterations.

In this case, the signal may be weaker than what should be, due to too many missing values (67 out of 170).

### Appendix V: Immunization

We selected 70 features related to immunization, which can be found in the feature list for immunization (supplemental materials). The results are:

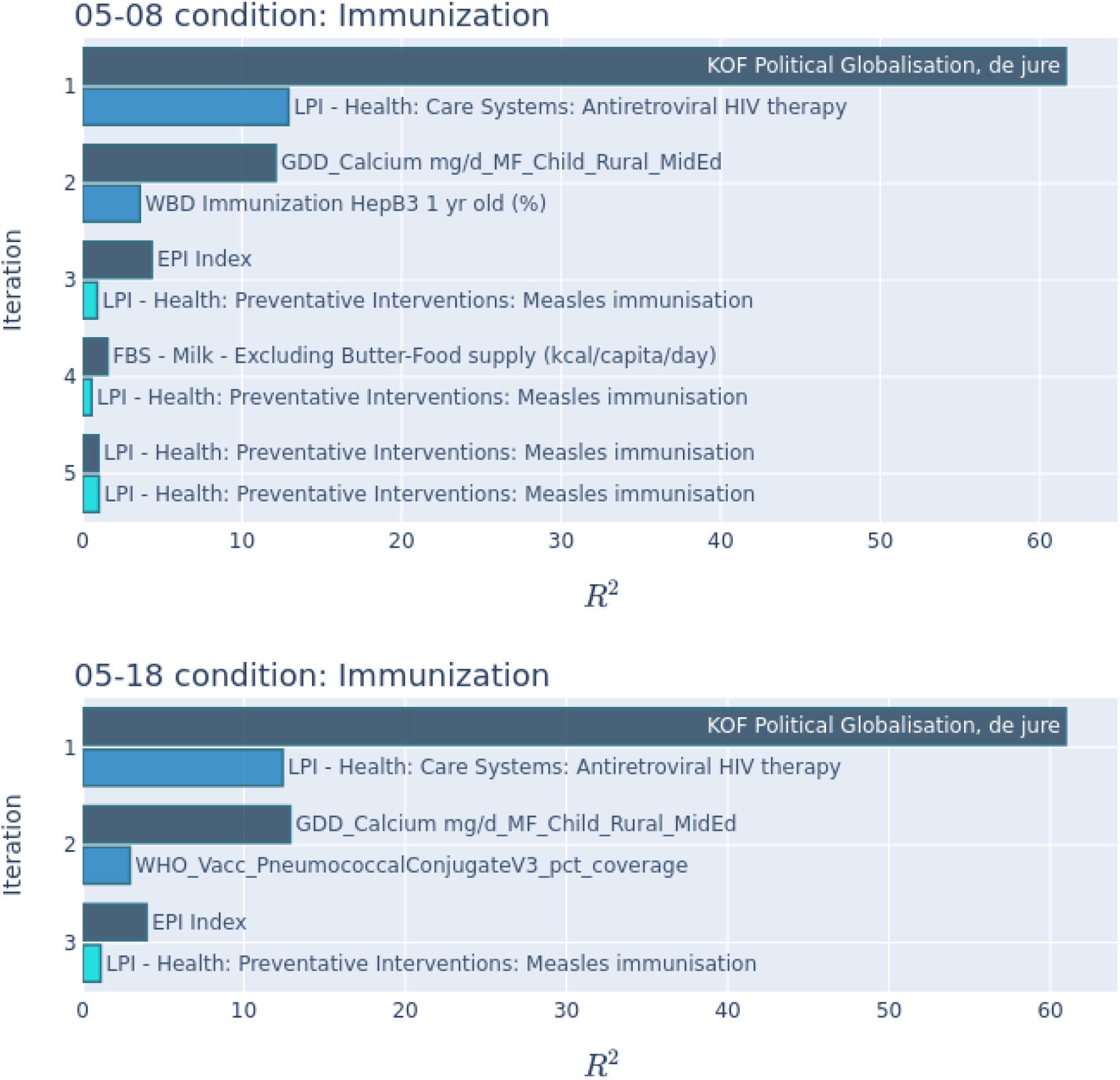

We do not find any of the contributions of immunization features significant. Both their absolute and relative *R*^2^are very small. Measles immunization appears consistently, so if there was any immunization that had an effect at the start of the pandemic, it was measles. Additionally, BCG immunization was thought to be important (46-49), but this cannot be proven with this data.

### Appendix VI: Inequality

We selected 9 features with inequality information, which can be found in the feature list for inequality (supplemental materials). The results are:

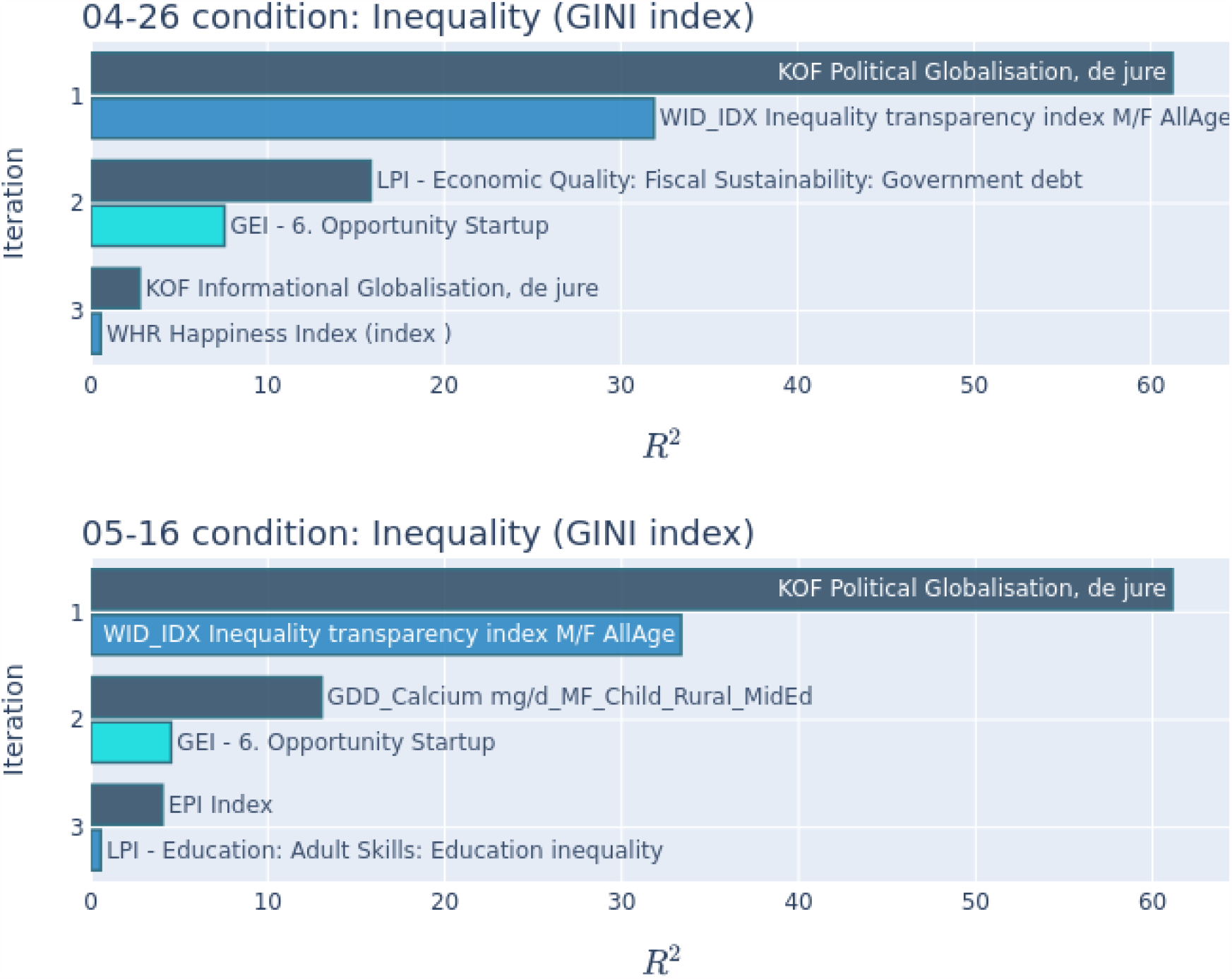

The inequality transparency index is half or less of the globalization for the first iteration. Since it does not appear later, it may be simply correlated to globalization. The other dominant feature, GEI Opportunity Startup, is not significant compared to the top ones of the group. No other feature in this group is significant. The Gini inequality index (50-51) does not appear to be significant at the onset of the pandemic.

### Appendix VII: Preventative Measures

Here we analyze the non-pharmaceutical preventative measures taken by different countries as a condition. Since these are usually trade-offs between costs and effectiveness, and can be easily decided by a government, there is a lot of interest in evaluating their potential (52-56).

We selected 228 features with preventative measures information, mostly extracted from the Oxford database (57), which can be found in the feature list for preventative measures (supplemental materials). The results are:

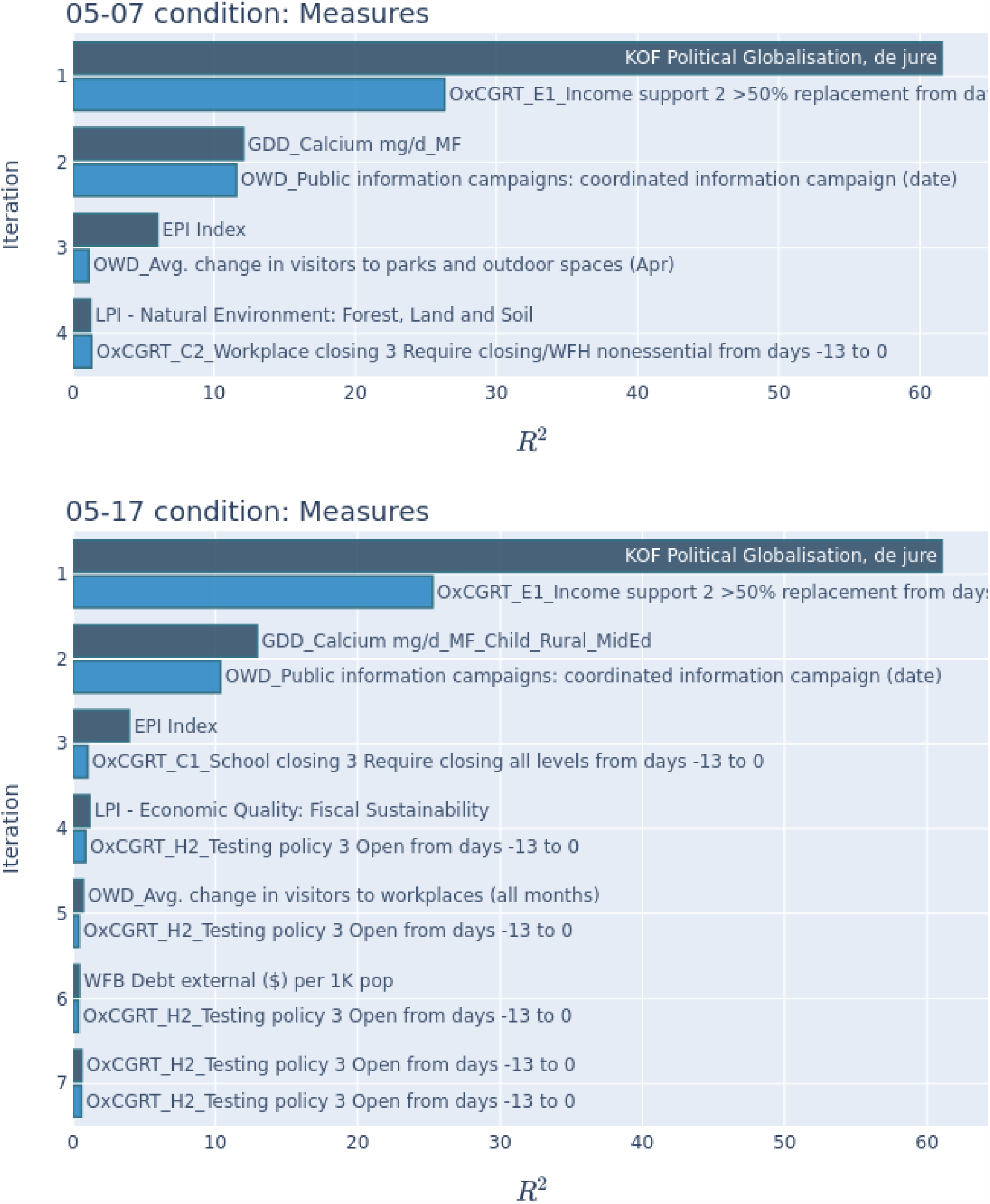

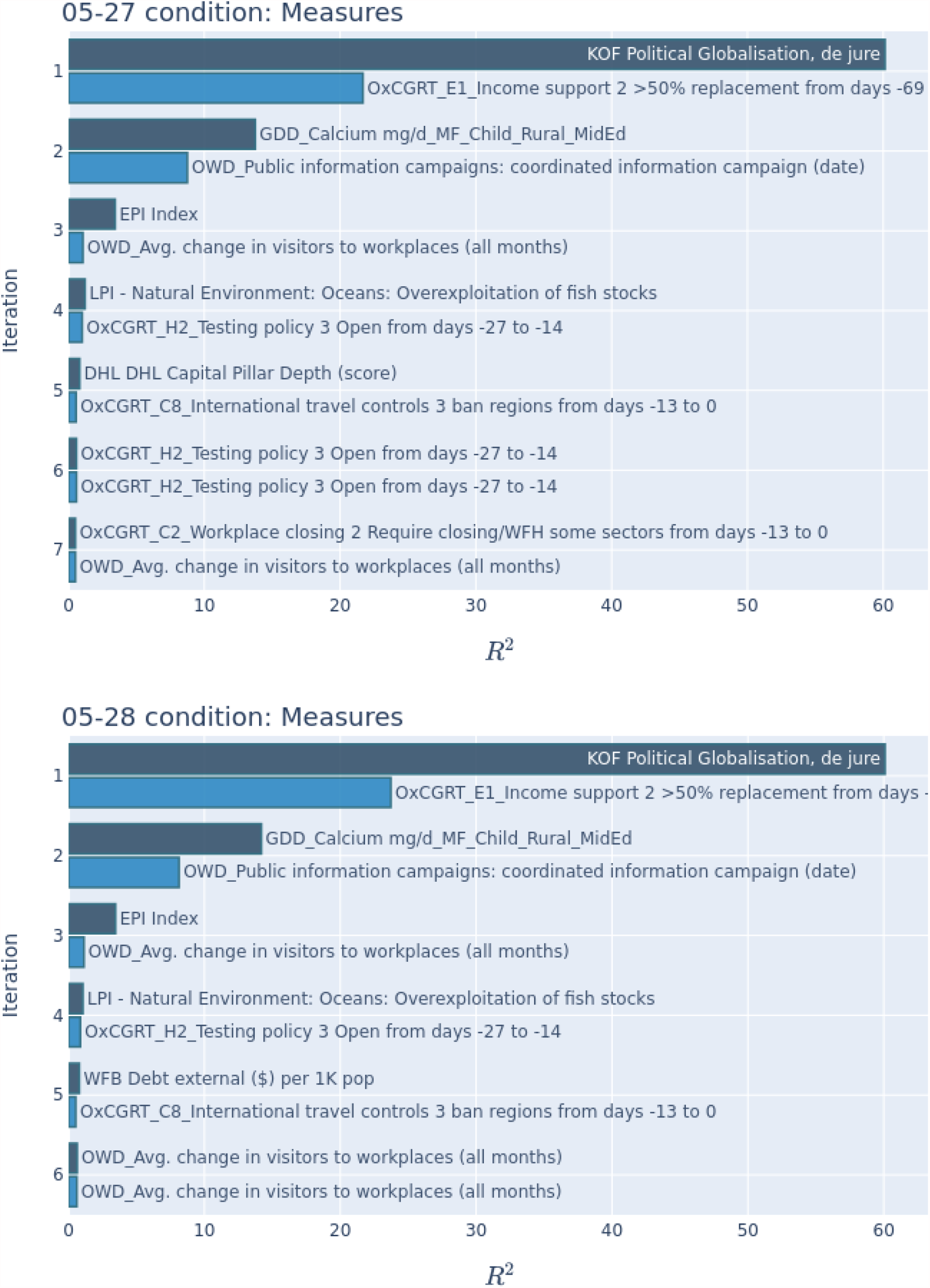

The first iteration shows a consistent feature, one of the forms of income support (at only ⅓ of the importance though). Since this feature disappears when we include globalization, it is an indication that income was a proxy for it (possibly correlated with wealth). This is another illustration of the power of Orthogonalization. On the second iteration we find a dominant measure, the public information campaigns, which at some point (05-07) are competitive with the selected feature. Change in visitors to workplaces also appears often, and in one case (05-28) is the top selected feature for the 6th iteration. Same with testing policy 3, on 05-17 and 27. This is a very promising analysis for later dates.

## Footnotes

1 We use the term feature from ML to mean the same as attributes from databases, independent variables from statistics, or columns from the matrix representation. We use the term target fro ML as dependent variable from statistics.

## Notes

### Competing Interest Statement

The authors have declared no competing interest.

### Funding Statement

No external funding was received.

### Author Declarations

No IRB approval is necessary as the research is only concerned with existing publicly available data.

## References

1. Rice, J. R. (1966). Experiments on gram-schmidt orthogonalization. Mathematics of Computation, 20(94), 325–328.

2. Shapley, L. S. (1953). A value for n-person games. Contributions to the Theory of Games, 2(28), 307–317.

3. Breiman, L. (2001). Random forests. Machine learning, 45(1), 5–32.

4. Fisher, A., Rudin, C., & Dominici, F. (2018). Model class reliance: Variable importance measures for any machine learning model class, from the” rashomon” perspective. arXiv preprint arXiv:1801.01489, 68.

5. Altmann, A., Toloşi, L., Sander, O., & Lengauer, T. (2010). Permutation importance: a corrected feature importance measure. Bioinformatics, 26(10), 1340–1347.

6. Saltelli, A. (2002). Sensitivity analysis for importance assessment. Risk analysis, 22(3), 579–590.

7. Saltelli, A., Ratto, M., Andres, T., Campolongo, F., Cariboni, J., Gatelli, D., … & Tarantola, S. (2008). Global sensitivity analysis: the primer. John Wiley & Sons.

8. Saltelli, A., Tarantola, S., Campolongo, F., & Ratto, M. (2004). Sensitivity analysis in practice: a guide to assessing scientific models (Vol. 1). New York: Wiley.

9. Fang, L., Karakiulakis, G., & Roth, M. (2020). Are patients with hypertension and diabetes mellitus at increased risk for COVID-19 infection?. The Lancet. Respiratory Medicine, 8(4), e21.

10. Gupta, R., Hussain, A., & Misra, A. (2020). Diabetes and COVID-19: evidence, current status and unanswered research questions. European journal of clinical nutrition, 74(6), 864–870.

11. Guo, W., Li, M., Dong, Y., Zhou, H., Zhang, Z., Tian, C., … & Hu, D. (2020). Diabetes is a risk factor for the progression and prognosis of COVID-19. Diabetes/metabolism research and reviews, 36(7), e3319.

12. Caussy, C., Pattou, F., Wallet, F., Simon, C., Chalopin, S., Telliam, C., … & Disse, E. (2020). Prevalence of obesity among adult inpatients with COVID-19 in France. The Lancet Diabetes & Endocrinology, 8(7), 562–564.

13. Stefan, N., Birkenfeld, A. L., Schulze, M. B., & Ludwig, D. S. (2020). Obesity and impaired metabolic health in patients with COVID-19. Nature Reviews Endocrinology, 16(7), 341–342.

14. Nakeshbandi, M., Maini, R., Daniel, P., Rosengarten, S., Parmar, P., Wilson, C., … & Breitman, I. (2020). The impact of obesity on COVID-19 complications: a retrospective cohort study. International Journal of Obesity, 44(9), 1832–1837.

15. Kang, Z., Luo, S., Gui, Y., Zhou, H., Zhang, Z., Tian, C., … & Hu, D. (2020). Obesity is a potential risk factor contributing to clinical manifestations of COVID-19. International Journal of Obesity, 44(12), 2479–2485.

16. Davies, N. G., Klepac, P., Liu, Y., Prem, K., Jit, M., & Eggo, R. M. (2020). Age-dependent effects in the transmission and control of COVID-19 epidemics. Nature medicine, 26(8), 1205–1211.

17. Cheng, Y., Luo, R., Wang, K., Zhang, M., Wang, Z., Dong, L., … & Xu, G. (2020). Kidney disease is associated with in-hospital death of patients with COVID-19. Kidney international, 97(5), 829–838.

18. Hirsch, J. S., Ng, J. H., Ross, D. W., Sharma, P., Shah, H. H., Barnett, R. L., … & Northwell COVID-19 Research Consortium. (2020). Acute kidney injury in patients hospitalized with COVID-19. Kidney international, 98(1), 209–218.

19. Zhang, L. K., Sun, Y., Zeng, H., Wang, Q., Jiang, X., Shang, W. J., … & Xiao, G. (2020). Calcium channel blocker amlodipine besylate therapy is associated with reduced case fatality rate of COVID-19 patients with hypertension. Cell discovery, 6(1), 1–12.

20. Allen, D. M. (1974). The relationship between variable selection

21. and data augmentation and a method for prediction. technometrics, 16(1), 125–127.

22. Stone, M. (1974). Cross-validatory choice and assessment of statistical predictions. Journal of the Royal Statistical Society: Series B (Methodological), 36(2), 111–133.

23. Stone, M. (1977). An asymptotic equivalence of choice of model by cross-validation and Akaike’s criterion. Journal of the Royal Statistical Society: Series B (Methodological), 39(1), 44–47.

24. Zietz, M., Zucker, J., & Tatonetti, N. P. (2020). Associations between blood type and COVID-19 infection, intubation, and death. Nature communications, 11(1), 1–6.

25. Lipsitch, M., Tchetgen, E. T., & Cohen, T. (2010). Negative controls: a tool for detecting confounding and bias in observational studies. Epidemiology (Cambridge, Mass.), 21(3), 383.

26. Jordan, R. E., Adab, P., & Cheng, K. (2020). Covid-19: risk factors for severe disease and death.

27. Zheng, Z., Peng, F., Xu, B., Zhao, J., Liu, H., Peng, J., … & Tang, W. (2020). Risk factors of critical & mortal COVID-19 cases: A systematic literature review and meta-analysis. Journal of Infection.

28. Li, X., Xu, S., Yu, M., Wang, K., Tao, Y., Zhou, Y., … & Zhao, J. (2020). Risk factors for severity and mortality in adult COVID-19 inpatients in Wuhan. Journal of Allergy and Clinical Immunology, 146(1), 110–118.

29. Zhou, F., Yu, T., Du, R., Fan, G., Liu, Y., Liu, Z., … & Cao, B. (2020). Clinical course and risk factors for mortality of adult inpatients with COVID-19 in Wuhan, China: a retrospective cohort study. The Lancet, 395(10229), 1054–1062.

30. Yang, J., Zheng, Y., Gou, X., Pu, K., Chen, Z., Guo, Q., … & Zhou, Y. (2020). Prevalence of comorbidities in the novel Wuhan coronavirus (COVID-19) infection: a systematic review and meta-analysis. Int J Infect Dis, 10.

31. Zhang, J. J., Dong, X., Cao, Y. Y., Yuan, Y. D., Yang, Y. B., Yan, Y. Q., … & Gao, Y. D. (2020). Clinical characteristics of 140 patients infected with SARS-CoV-2 in Wuhan, China. Allergy, 75(7), 1730–1741.

32. Huang, C., Wang, Y., Li, X., Ren, L., Zhao, J., Hu, Y., … & Cao, B. (2020). Clinical features of patients infected with 2019 novel coronavirus in Wuhan, China. The Lancet, 395(10223), 497–506.

33. Chen, N., Zhou, M., Dong, X., Qu, J., Gong, F., Han, Y., … & Zhang, L. (2020). Epidemiological and clinical characteristics of 99 cases of 2019 novel coronavirus pneumonia in Wuhan, China: a descriptive study. The Lancet, 395(10223), 507–513.

34. Chen, T., Wu, D. I., Chen, H., Yan, W., Yang, D., Chen, G., … & Ning, Q. (2020). Clinical characteristics of 113 deceased patients with coronavirus disease 2019: retrospective study. bmj, 368.

35. Zhu, L., She, Z. G., Cheng, X., Qin, J. J., Zhang, X. J., Cai, J., … & Li, H. (2020). Association of blood glucose control and outcomes in patients with COVID-19 and pre-existing type 2 diabetes. Cell metabolism, 31(6), 1068–1077.

36. Gkisser, S. (2017). Predictive inference: an introduction. Chapman and Hall/CRC.

37. Kohavi, R. (1995, August). A study of cross-validation and bootstrap for accuracy estimation and model selection. In Ijcai (Vol. 14, No. 2, pp. 1137–1145).

38. Gygli, S., Haelg, F., Potrafke, N., & Sturm, J. E. (2019). The KOF globalisation index–revisited. The Review of International Organizations, 14(3), 543–574.

39. Miller, V., Singh, G. M., Onopa, J., Reedy, J., Shi, P., Zhang, J., … & Mozaffarian, D. (2021). Global Dietary Database 2017: data availability and gaps on 54 major foods, beverages and nutrients among 5.6 million children and adults from 1220 surveys worldwide. BMJ global health, 6(2), e003585.

40. Index, E. P. (2018). Environmental performance index. Yale University and Columbia University: New Haven, CT, USA.

41. Covid-19 Coronavirus Pandemic. Worldometer. (2021, June 3). https://www.worldometers.info/coronavirus/.

42. Dong, E., Du, H., & Gardner, L. (2020). An interactive web-based dashboard to track COVID-19 in real time. The Lancet infectious diseases, 20(5), 533–534.

43. Baud, D., Qi, X., Nielsen-Saines, K., Musso, D., Pomar, L., & Favre, G. (2020). Real estimates of mortality following COVID-19 infection. The Lancet infectious diseases, 20(7), 773.

44. Latz, C. A., DeCarlo, C., Boitano, L., Png, C. M., Patell, R., Conrad, M. F., … & Dua, A. (2020). Blood type and outcomes in patients with COVID-19. Annals of hematology, 99(9), 2113–2118.

45. Rubin, R. (2020). Investigating whether blood type is linked to COVID-19 risk. Jama.

46. Gross, S. (1980, August). Median estimation in sample surveys. In Proceedings of the Section on Survey Research Methods (Vol. 1814184). Alexandria, VA: American Statistical Association.

47. Pawlowski, C., Puranik, A., Bandi, H., Venkatakrishnan, A. J., Agarwal, V., Kennedy, R., … & Soundararajan, V. (2021). Exploratory analysis of immunization records highlights decreased SARS-CoV-2 rates in individuals with recent non-COVID-19 vaccinations. Scientific reports, 11(1), 1–20.

48. Redelman-Sidi, G. (2020). Could BCG be used to protect against COVID-19?. Nature Reviews Urology, 17(6), 316–317.

49. Curtis, N., Sparrow, A., Ghebreyesus, T. A., & Netea, M. G. (2020). Considering BCG vaccination to reduce the impact of COVID-19. The Lancet, 395(10236), 1545–1546.

50. O’Neill, L. A., & Netea, M. G. (2020). BCG-induced trained immunity: can it offer protection against COVID-19?. Nature Reviews Immunology, 20(6), 335–337.

51. Ahmed, F., Ahmed, N. E., Pissarides, C., & Stiglitz, J. (2020). Why inequality could spread COVID-19. The Lancet Public Health, 5(5), e240.

52. Oronce, C. I. A., Scannell, C. A., Kawachi, I., & Tsugawa, Y. (2020). Association between state-level income inequality and COVID-19 cases and mortality in the USA. Journal of General Internal Medicine, 35(9), 2791–2793.

53. Ferguson, N. M., Laydon, D., Nedjati-Gilani, G., Imai, N., Ainslie, K., Baguelin, M., … & Van-Elsland, S. (2020). Impact of non-pharmaceutical interventions (NPIs) to reduce COVID-19 mortality and healthcare demand. Imperial College COVID-19 Response Team. Imperial College COVID-19 Response Team, 20.

54. Vardavas, R., de Lima, P. N., & Baker, L. (2021). Modeling COVID-19 Nonpharmaceutical Interventions: Exploring periodic NPI strategies. medRxiv.

55. Brauner, J. M., Mindermann, S., Sharma, M., Johnston, D., Salvatier, J., Gavenčiak, T., … & Kulveit, J. (2021). Inferring the effectiveness of government interventions against COVID-19. Science, 371(6531).

56. Bo, Y., Guo, C., Lin, C., Zeng, Y., Li, H. B., Zhang, Y., … & Lao, X. Q. (2021). Effectiveness of non-pharmaceutical interventions on COVID-19 transmission in 190 countries from 23 January to 13 April 2020. International Journal of Infectious Diseases, 102, 247–253.

57. Duhon, J., Bragazzi, N., & Kong, J. D. (2021). The impact of non-pharmaceutical interventions, demographic, social, and climatic factors on the initial growth rate of COVID-19: A cross-country study. Science of The Total Environment, 760, 144325.

58. Hale, T., Webster, S., Petherick, A., Phillips, T., & Kira, B. (2020). Oxford COVID-19 government response tracker (OxCGRT).

