## Supplementary material for "Analysis of feature influence on Covid-19 Death Rate Per Country Using a Novel Orthogonalization Technique": Selected features by iteration

| Date | rows | cols | Iter 1 | Feature | Iter 2 | Feature | Iter 3 | Feature | Iter 4 | Feature | Iter 5 | Feature | Iter 6 | Feature | Iter 7 | Feature | Iter 8 | Feature |
|---|---|---|---|---|---|---|---|---|---|---|---|---|---|---|---|---|---|---|
| 2020-04-15 | 165 | 3217 | 62.54 | Political Globalisation | 18.09 | Climate preparedness index | 2.45 | Fruits | 1.57 | EPI Index | 0.60 | Capital Pillar Breadth | 0.44 | Telephones fixed lines | 0.12 | Utility model applications | 0.17 | Stay-at-home restrictions |
| 2020-04-16 | 165 | 3217 | 60.30 | Political Globalisation | 19.69 | LPI Material Resources | 4.31 | Cocoa Beans and products | 0.61 | Vegetable supply | 0.21 | Labor force | 0.16 | Poverty Gap health expends | 0.13 | Capital Pillar | 0.15 | Citrus, Other |
| 2020-04-17 | 165 | 3217 | 61.56 | Political Globalisation | 20.40 | LPI Regulatory quality | 2.18 | LPI Waged workers | 1.32 | Capital Pillar Breadth | 0.43 | Blood O neg % | 0.49 | Fish, Liver Oil | 0.16 | Citrus, Other | 0.19 | Infant mortality rate |
| 2020-04-18 | 165 | 3218 | 60.24 | Political Globalisation | 18.95 | LPI Regulatory quality | 2.62 | Blood O neg % | 1.22 | OWD Debt and contract relief | 0.90 | Capital Pillar Depth | 0.60 | Coconut Oil | 0.27 | Deaths chronic resp disease | 0.24 | Natural gas production |
| 2020-04-25 | 165 | 3223 | 61.80 | Political Globalisation | 15.55 | LPI Government debt | 3.50 | Capital Pillar Breadth | 0.96 | GDD Calcium | 0.76 | Capital Pillar Depth | 0.64 | Refugee population | 0.45 | Telephones fixed lines | 0.14 | Ethnic SubSaharan |
| 2020-04-26 | 165 | 3223 | 61.22 | Political Globalisation | 15.84 | LPI Government debt | 2.77 | Informational Globalisation | 2.30 | GDD Calcium | 0.49 | cellular prices | 0.36 | Global Connectedness Depth | 0.30 | Health expenditure | 0.32 | Air transport departures |
| 2020-04-27 | 165 | 3223 | 60.83 | Political Globalisation | 15.39 | LPI Government debt | 2.73 | Capital Pillar Breadth | 1.00 | GDD Calcium | 0.98 | Capital Pillar Depth | 0.62 | Telephones fixed lines | 0.69 | Foreign direct investment | 0.35 | Security of women |
| 2020-04-28 | 165 | 3223 | 60.35 | Political Globalisation | 15.94 | LPI Government debt | 2.84 | Refined petro product | 1.10 | UN region 155 | 0.96 | Foreign direct investment | 2.76 | Capital Pillar Depth | 0.69 | Death Parkinson disease | 0.32 | Crude oil production |
| 2020-05-05 | 167 | 3223 | 62.59 | Political Globalisation | 12.13 | GDD Calcium | 5.65 | EPI Index | 1.28 | Capital Pillar Depth | 0.55 | LPI Measles immunisation | 0.54 | Intl travel controls 1 | 0.52 | Provincial hospitals | 0.24 | Foreign direct investment |
| 2020-05-06 | 167 | 3223 | 61.93 | Political Globalisation | 12.02 | GDD Calcium | 6.09 | EPI Index | 1.35 | Capital Pillar Depth | 1.29 | Research and development | 0.52 | Provincial hospitals | 0.25 | Population 55 to 64 F | 0.17 | Vacc Rotavirus1 |
| 2020-05-07 | 167 | 3224 | 61.57 | Political Globalisation | 12.07 | GDD Calcium | 5.99 | EPI Index | 1.25 | LPI Forest, Land and Soil | 0.87 | LPI Health | 0.48 | Population 55 to 64 F | 0.28 | ef regulation credit | 0.27 | Testing policy 3 |
| 2020-05-08 | 167 | 3225 | 61.63 | Political Globalisation | 12.11 | GDD Calcium | 4.34 | EPI Index | 1.56 | Milk - Excluding Butter | 1.01 | LPI Measles immunisation | 0.60 | Health expenditure | 0.30 | WFB Debt external | 0.24 | Testing policy 3 |
| 2020-05-15 | 168 | 3225 | 61.20 | Political Globalisation | 12.86 | GDD Calcium | 3.95 | EPI Index | 1.19 | LPI Fiscal Sustainability | 0.74 | Change in visits to workpl | 0.64 | Testing policy 3 | 0.61 | temperate climate | 0.43 | Blood A pos |
| 2020-05-16 | 168 | 3225 | 61.12 | Political Globalisation | 13.04 | GDD Calcium | 4.05 | EPI Index | 1.12 | LPI Fiscal Sustainability | 0.75 | Change in visits to workpl | 0.44 | Intnl travel screening | 0.44 | WFB Debt external | 0.36 | Getting Electricity |
| 2020-05-17 | 168 | 3225 | 61.05 | Political Globalisation | 12.93 | GDD Calcium | 3.95 | EPI Index | 1.18 | LPI Fiscal Sustainability | 0.72 | Change in visits to workpl | 0.44 | WFB Debt external | 0.62 | Testing policy 3 | 0.24 | NOAA DecELEVATION |
| 2020-05-18 | 168 | 3225 | 60.96 | Political Globalisation | 12.88 | GDD Calcium | 3.97 | EPI Index | 1.14 | LPI Fiscal Sustainability | 0.75 | Change in visits to workpl | 0.47 | Intnl travel screening | 0.44 | Capital Pillar Depth | 0.37 | WFB Debt external |
| 2020-05-25 | 168 | 3225 | 60.42 | Political Globalisation | 13.83 | GDD Calcium | 3.64 | EPI Index | 1.14 | LPI Overexploitation of fish | 0.77 | Capital Pillar Depth | 0.52 | Testing policy 3 | 0.44 | Change in visits to workpl | 0.45 | Budget surplus |
| 2020-05-26 | 168 | 3225 | 60.47 | Political Globalisation | 14.19 | GDD Calcium | 3.58 | EPI Index | 1.13 | LPI Overexploitation of fish | 0.82 | WFB Debt external | 0.61 | Change in visits to workpl | 0.40 | Testing policy 3 | 0.47 | Getting Electricity |
| 2020-05-27 | 168 | 3225 | 60.13 | Political Globalisation | 13.75 | GDD Calcium | 3.42 | EPI Index | 1.21 | LPI Overexploitation of fish | 0.83 | Capital Pillar Depth | 0.58 | Testing policy 3 | 0.49 | Workplace closing 2 | 0.44 | Restr internal movement 2 |
| 2020-05-28 | 168 | 3225 | 60.09 | Political Globalisation | 14.19 | GDD Calcium | 3.44 | EPI Index | 1.05 | LPI Overexploitation of fish | 0.78 | WFB Debt external | 0.63 | Change in visits to workpl | 0.37 | Testing policy 3 | 0.39 | Getting Electricity |
